# Possible explanations for the discrepancy between randomised trials and observational studies of the effectiveness of 4CMenB vaccine against gonorrhoea: a modelling study

**DOI:** 10.64898/2026.11.27.26369132

**Authors:** Jørgen E. Midtbø, Gunnar Rø, Ola Brynildsrud, Vegard Eldholm, Jon Michael Gran, Marissa LeBlanc

## Abstract

**Background:** Evidence from observational studies suggests that the 4CMenB vaccine offers cross-protection against gonorrhoea. On this basis, England implemented a targeted national 4CMenB programme, but recent randomised controlled trials (RCTs) have found little evidence of protection. Proposed explanations include confounding of the observational studies and biological mechanisms; we investigated the latter.

**Methods:** Using a transmission model of gonorrhoea and chlamydia, we simulated both an RCT and a case-control study within high- and low-risk subpopulations, respectively, in the same epidemic. Our simulations allowed for two mechanisms, blunting of vaccine response by recent gonorrhoea and heterogeneity in exposure intensity due to different sexual behaviour. RCT and case-control estimates from simulated data were compared to real-world findings. Additional simulations were performed with a programme like England’s.

**Results:** In simulations with a truly protective vaccine, differences in key population characteristics combined with the mechanisms above were sufficient to simultaneously replicate both real-world RCT and case-control estimates in respective subpopulations. In simulations with a vaccine programme, diagnoses among targeted individuals fell by 30–46% in year 3 (15–57% across assumptions) compared to no programme, mostly through indirect protection.

**Conclusions:** Our study shows that the discrepancy between trial and observational estimates of vaccine effectiveness can be plausibly explained by biological mechanisms. Thus, 4CMenB may protect, but less in recently infected or intensely exposed individuals whom programmes target. Stratified analyses of protection could shed light on the mechanisms.

## 1 Introduction

Gonorrhoea is a sexually transmitted infection (STI) caused by *Neisseria gonorrhoeae* (NG), which infects the urethra, cervix, rectum and pharynx. Infections are often asymptomatic and can lead to pelvic inflammatory disease, ectopic pregnancy and infertility,^1^ and antimicrobial resistance is a growing challenge.^2^

No vaccine against NG exists,^3^ but meningococcal group B vaccines with outer membrane vesicles (OMV), which carry proteins shared with NG, may give cross-protection. Observational studies have estimated vaccine effectiveness (VE) against NG at 30–45%,^4–9^ and in August 2025 England began offering the four-component vaccine 4CMenB (Bexsero) to gay, bisexual, and other men who have sex with men (GBMSM) at high risk.^10^

However, four randomised controlled trials (RCTs) in GBMSM have either been inconclusive or found little or no protection: the open-label DOXYVAC in France (adjusted hazard ratio 0.78, 95% CI 0.60–1.01),^11^ MenGO in Australia (incidence rate ratio [IRR] 0.78, 0.40–1.51),^12^ the placebo-controlled trial in Hong Kong (hazard ratio 0.76, 0.31–1.87)^13^ and GoGoVax in Australia (IRR 1.01, 0.80–1.26; VE 0.5%).^14^ Only GoGoVax, the largest, had a confidence interval that excluded a VE above 0.20. An editorial in *The Lancet Infectious Diseases* concluded that GoGoVax dispels hopes for gonorrhoea vaccination.^15^

The discrepancy between the RCTs and observational studies may reflect differences in estimands,^16^ confounding^14^ or collider bias in the observational studies,^17^ although a similar observational study on the effects of meningococcal group B vaccine without OMV showed no effect, arguing against healthy-vaccinee bias.^18^ Two biological mechanisms could also make the same vaccine protect less in the trials’ participants.

First, prior infection may blunt the vaccine response. In MenGO, bactericidal (hSBA) titres did not rise in participants with NG at a vaccine dose,^12^ although hSBA is not an established correlate of protection against NG. Antibodies to the NG outer-membrane reduction-modifiable protein (Rmp), induced by infection and possibly boosted by its meningococcal homologue in 4CMenB, block complement-mediated killing by other antibodies,^19^ and GoGoVax’s investigators suggested that such blunting may explain their result.^14^ How strong blunting might be, and how recent an infection must be to cause it, is unknown.

Second, for a leaky vaccine (*i.e.* a vaccine that reduces rather than eliminates susceptibility), breakthrough infection depends not only on how often one is exposed but on the intensity of those exposures. Indeed, modelling of malaria and HIV prevention trials has shown that the measured efficacy of a leaky intervention is reduced under intense or uneven exposure.^20,21^ Sex involving several anatomical sites or many partners could expose the RCTs’ participants, selected for high risk and in GoGoVax mostly for recent NG, more intensely than the observational studies’ populations. Such heterogeneity in exposure intensity has not been tested as an explanation for the discrepancy.

We therefore used a transmission model to test whether these two mechanisms can reconcile the trial and observational estimates, by simulating an RCT and an observational study of the same vaccine in the same epidemic, and what they would mean for a programme like England’s.

## 2 Methods

### 2.1 Study design

We built an agent-based model of transmission of NG and chlamydia (CT), with a structure similar to previous network-based gonorrhoea models.^22^ It simulated two linked populations, representing (1) GBMSM and (2) majority-heterosexual adolescents and young adults. In every simulation, we conducted a randomised trial modelled on GoGoVax in the first population and a case-control study with CT controls, the most common observational design, in the second, so that both estimated the VE of the same vaccine in the same epidemic (Figure 1). We varied assumptions about sexual networks, transmission and the vaccine, and identified the combinations that reproduced both GoGoVax’s and the observational studies’ estimates. The model was set up with plausible parameters, but kept deliberately simple, as the aim was to test feasibility of the hypotheses, not to make quantitative predictions. The study used no individual-level data and did not require ethical approval.

**Figure 1:**
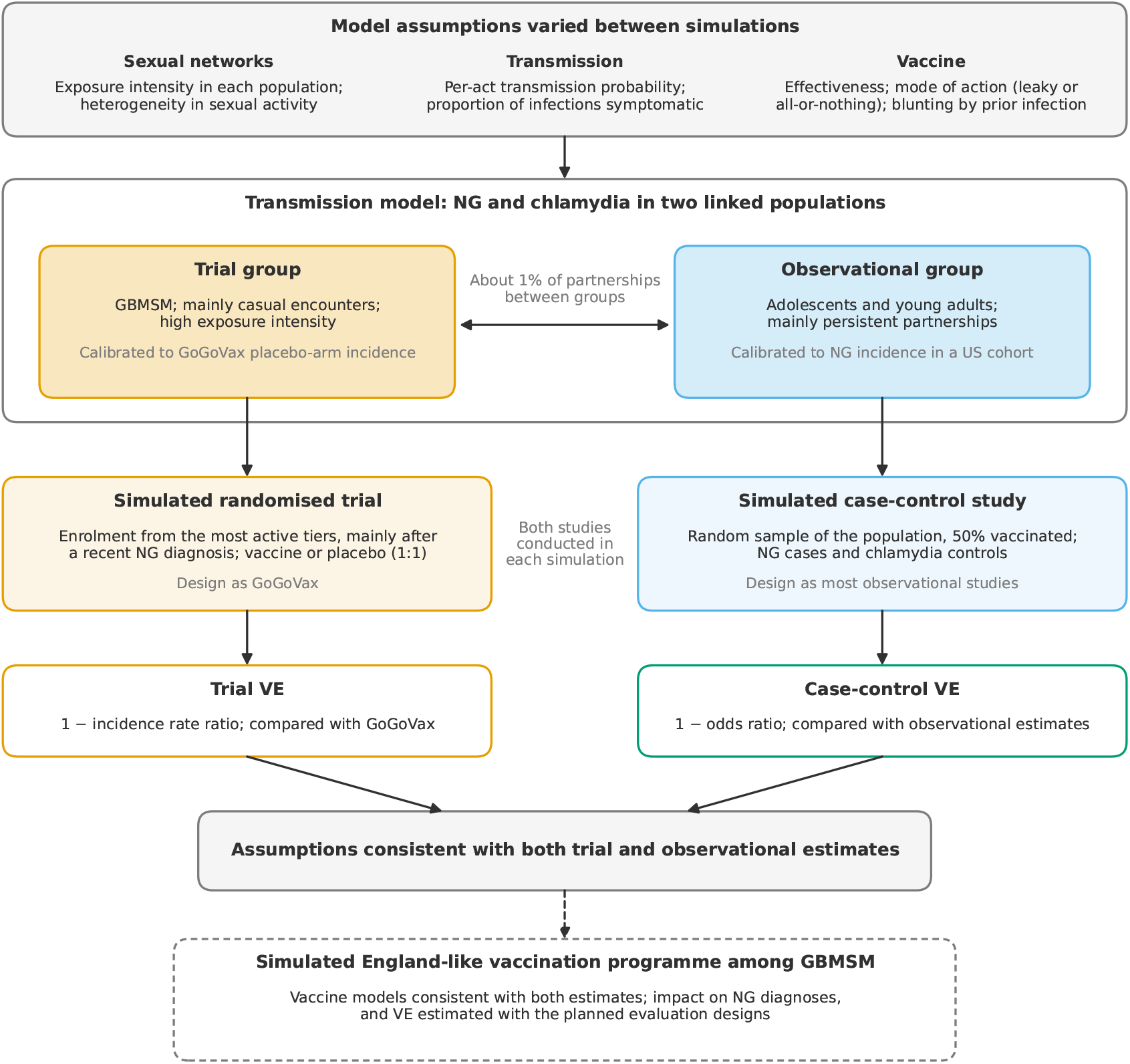
Study design. Each simulation models NG and chlamydia transmission in two linked populations, representing GBMSM (trial group) and adolescents and young adults (observational group), under a common set of model assumptions. Incidences calibrated to GoGoVax^14^ and a US cohort,^6^ respectively. In each simulation, a randomised trial with GoGoVax’s design is conducted in the trial group and a case-control study with chlamydia controls in the observational group, so that both estimate the VE of the same vaccine. Vaccine models consistent with both GoGoVax’s and the observational studies’ estimates are then used to simulate an England-like programme. GBMSM: gay, bisexual, and other men who have sex with men.

### 2.2 Simulated populations

The two populations, hereafter the trial and observational groups, each comprised 100,000 sexually active people, with about 1% of partnerships between groups. Each group was divided into four tiers of sexual activity holding 60%, 25%, 10% and 5% of its members (Supplementary Figure S1); the trial group’s two highest tiers stood for GBMSM on PrEP or living with HIV. Partnerships were either persistent, lasting 6 months on average, or one-off casual encounters, and could be concurrent. In the trial group, 70% of acts were in casual encounters, with four acts per encounter on average to stand for several anatomical sites, and each act carried a transmission probability of 0.8 or 0.1 with equal likelihood, standing for high- and low-transmission acts and sites. In the observational group, 30% of acts were in casual encounters, with 1.5 acts per encounter.

We summarised each group’s exposure intensity by *P*^-^*e*, the incidence-weighted mean probability that an exposure episode infects an unvaccinated susceptible person (Supplement). An exposure episode is one casual encounter with an infectious partner, or the run of acts with an infectious persistent partner until the infection is cleared or the partnership ends. Because *P*^-^*_e_* is weighted by incidence, it describes the episodes that produce cases, from which VE is estimated.

We refer to each set of assumptions about the sexual networks as a model *world*, one per observational setting (below). In our main analysis, the differential-intensity worlds, the trial group’s exposure episodes were more likely to infect than the observational group’s (*P*^-^*_e_* 0.92 against 0.57), mainly because of its multi-act casual encounters. In sensitivity analyses we applied the observational group’s partnership structure and transmission parameters also to the trial group, so that the groups differed only in partner numbers and had the same *P*^-^*_e_* (equal-intensity worlds); added variation in activity within the trial group’s tiers (activity weights); or added low-risk encounters that raised its partner numbers to those reported by trial participants (light encounters; Supplement).

Heterosexual transmission parameters are poorly known, so results are pooled over three plausible settings of the observational group’s per-act transmission probability and symptomatic share of NG (Supplement). In the trial group, 10% of NG infections were symptomatic.

NG and CT were modelled as susceptible–infectious–susceptible infections transmitted over the same acts. People were tested at routine visits, with mean intervals from 24 months in the least active tier to 3 months in the most active, and when symptomatic. Every test detected all current NG and CT infections and all detected infections were successfully treated. Each group’s partnership rates were calibrated to its NG incidence, about 45 first diagnoses per 100 person-years in the simulated trial’s placebo arm (GoGoVax, 47.8^14^) and 0.52 among the observational group’s unvaccinated,^6^ and its CT per-act probability to the ratio of CT to NG (Supplement). Each simulation had a 25-year burn-in, to stabilise transmission dynamics, before the studies started.

### 2.3 Vaccine protection and the two hypotheses

The vaccine acted only on susceptibility to infection, from 28 days after the second dose, and did not wane (except in some programme simulations; below). Since it is not known how 4CMenB protects against NG, we considered the two standard modes of protection, with strength set by *V*_0_: An all-or-nothing vaccine gives a share *V*_0_ of vaccinees full protection and the rest none. A leaky vaccine instead lowers everyone’s hazard of infection by *V*_0_, which for a low risk of infection per exposure is approximately the relative reduction in that risk.

We tested whether two hypotheses could explain the discrepancy, alone or together. The first is blunting of the vaccine response by recent NG infection, set by a blunting window before dose 1 in which an NG infection blunts the response, or only by NG at time of vaccination, and a strength *b*, from no blunting (*b* = 1) to full blunting (*b* = 0), that multiplies *V*_0_. The second is heterogeneity in exposure intensity between the groups: a leaky vaccine protects less against exposure episodes that are more likely to infect^20,21^ (Supplement).

### 2.4 Simulated VE studies

On the day the burn-in ended, 650 people were drawn from the trial group’s two highest tiers, 90% of them among those diagnosed with NG in the previous 18 months and 10% among those not, standing for GoGoVax’s enrolment through syphilis, and randomly assigned 1:1 to vaccine or placebo.^14^ Doses were given on days 0 and 90. Follow-up started 28 days after dose 2 and lasted 730 days, with tests every 90 days and at symptomatic presentation and no loss to follow-up. The outcome was the first NG diagnosis.

On the same day, a random half of the observational group formed the case-control study’s cohort, and a random half of the cohort was vaccinated on the same schedule. Over the trial’s follow-up window, cases were cohort members whose first NG or CT diagnosis included NG, and controls those whose first such diagnosis was CT alone.

### 2.5 Statistical analysis

Trial VE was one minus the incidence rate ratio (IRR) of first NG diagnosis between the arms, as in GoGoVax,^14^ and case-control VE one minus the odds ratio (OR) of vaccination between cases and controls. Each vaccine model was run many times in each of the three settings and results pooled, in order to obtain precise estimates of the model’s expected prediction that could be compared against the uncertainty intervals of the real-world studies (Supplement).

### 2.6 Agreement criteria

We say that a vaccine model (mode of protection, *V*_0_ and blunting) agreed with the data if its case-control VE lay within the 95% CI of the pooled case-control estimate of a meta-analysis of the observational studies, 0.34 (0.27–0.41),^9^ and its trial VE within GoGoVax’s 68% CI ( 0.129 to 0.105). It was at the edge if its case-control VE was in that interval and its trial VE lay between GoGoVax’s 68% and 95% CIs (upper limit 0.20), and disagreed otherwise. We used the 68% CI as the main criterion because a vaccine model whose expected trial VE lies near the 95% limit reproduces GoGoVax’s estimate only if the GoGoVax estimate was an unusually low draw. An alternative way to quantify agreement is in the Supplement (Figure S5).

### 2.7 England-like programme

We simulated a programme like the one England has offered GBMSM since August 2025^23^ in the trial group, over three years. People were eligible for vaccination at any test or symptomatic visit if they were in the two highest tiers, standing for at least five partners in three months, or had been diagnosed with NG or CT in the past year, standing for a bacterial STI. A fixed 60% of people, drawn independently of risk, accepted vaccination, a share chosen to match the 41% of eligible GBMSM who reported a first dose by November–December 2025;^24^ acceptors received dose 1 at their first eligible visit and dose 2 at least 28 days later. We used vaccine models that agreed or came close for each hypothesis, with and without waning (mean duration of protection 3 years), and ran each with and without the programme from the same starting point: runs with the same random seed are identical until the programme starts and diverge after it. We report NG diagnoses in year 3 relative to no programme, and the two-dose VE that a GoGoVax-sized trial and UKHSA’s three planned designs would estimate: a cohort with vaccination as a time-varying exposure, a test-negative design and a case-control design with CT controls.^10^ Test-negative VE was also estimated within strata of NG at the vaccination visit, NG diagnosis in the previous year and route of eligibility, and the power to detect differences between strata by scaling the run-to-run variation to England’s number of GBMSM diagnoses (Supplement).

### 2.8 Tools and use of AI

The model and VE estimation code were written in Python with Claude Code (Opus 5.5; Anthropic) under close supervision (see Use of AI).

## 3 Results

The case-control VE constrained *V*_0_ rather than the mechanisms (Figure 2). A leaky vaccine gave a case-control VE of 0.30–0.33 with *V*_0_ = 0.5 and 0.38–0.42 with *V*_0_ = 0.6, both below *V*_0_ because the observational group’s exposure episodes are also fairly likely to infect (*P*^-^*_e_* 0.57). Blunting hardly moved it, because about 1% of observational vaccinees had been infected in the 18 months before vaccination, against 88% of trial enrollees: full blunting over 18 months lowered the case-control VE by 0.01–0.06 and the trial VE by 0.11–0.51 (Figure S4). The test of each hypothesis was thus whether it agreed with the trials.

**Figure 2:**
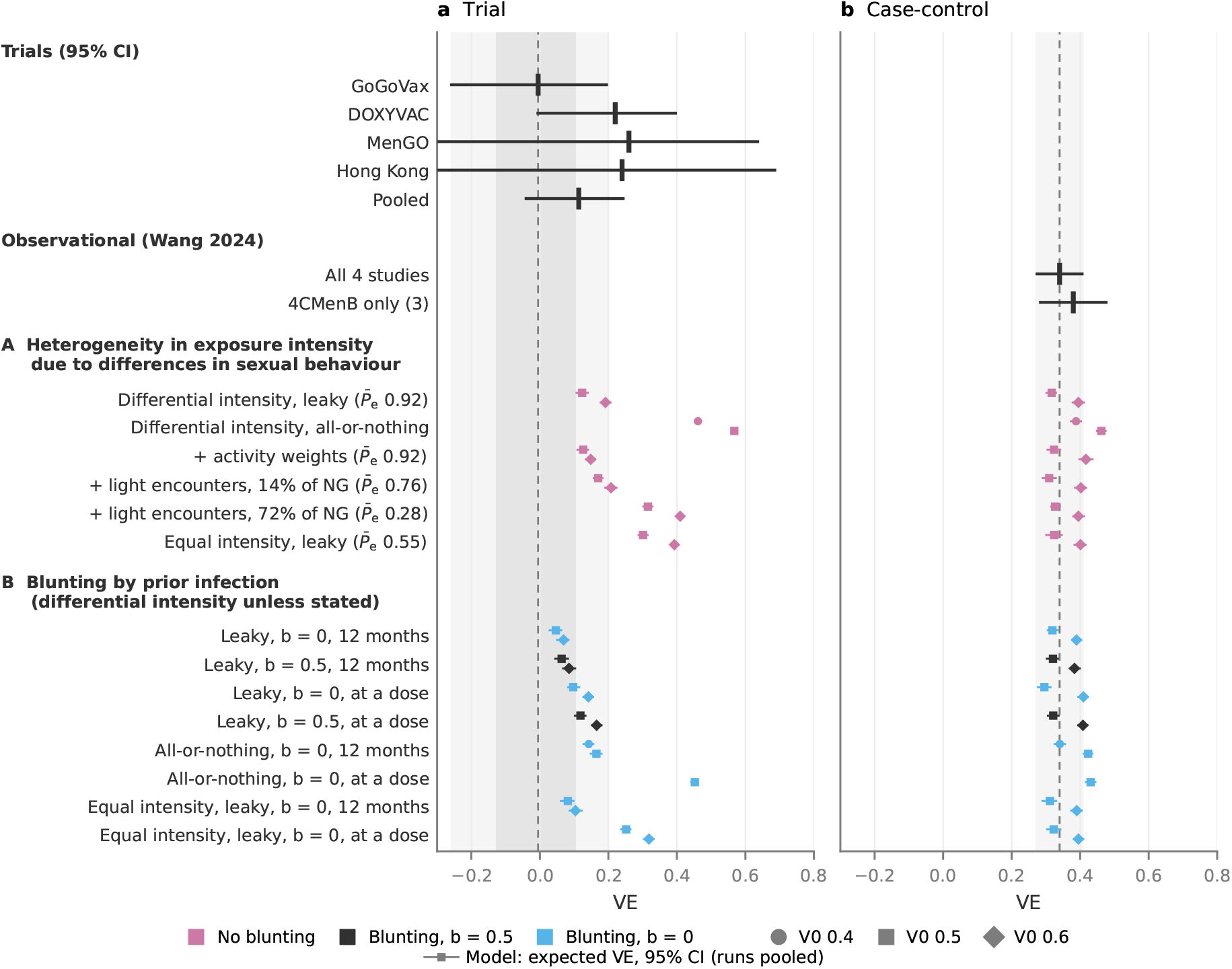
Estimated VE under each hypothesis in the trial (incidence rate ratio; a) and the observational case-control study (odds ratio; b), simulated together in the same runs and each pooled over many runs. Coloured markers show the model’s expected VE with a 95% CI that measures the uncertainty on the expected estimate, not on a single study; colour gives the blunting and marker shape the value of the intrinsic vaccine protection *V*_0_. In panel (a), the dashed line and shaded bands show GoGoVax’s point estimate and its 68% and 95% CIs.^14^ Black markers show data with 95% CIs: at the top, the four trials^11–14^ and our fixed-effect, inverse-variance pooling of their log rate or hazard ratios, 0.11 ( 0.04 to 0.25), shown for reference only; the agreement criteria use GoGoVax alone. In panel (b), the dashed line and shaded band show the pooled case-control estimate and its 95% CI, 0.34 (0.27–0.41),^9^ also shown as a black marker. (A) Heterogeneity in exposure intensity alone, without blunting: the differential-intensity worlds; sensitivity analyses that add to them, under leaky protection, activity weights, or activity weights and light encounters carrying 14% or 72% of the trial’s NG (“+” rows); and the equal-intensity worlds. The trial group’s *P*^-^*_e_*, the mean probability that an exposure episode infects, is given in brackets (see Methods for details). (B) Blunting by prior infection, in the differential-intensity worlds unless stated: full (*b* = 0) or half (*b* = 0.5), over the 12 months before dose 1 or by NG at a dose only. Vaccine protection is leaky unless stated. Other blunting windows are shown in Figure S3, and every vaccine model of the differential-intensity worlds in Figure S4.

Heterogeneity in exposure intensity alone brought a leaky vaccine to the edge of GoGoVax’s result. Without blunting, the trial VE was 0.12 (95% CI 0.10–0.14) with *V*_0_ = 0.5 and 0.19 with *V*_0_ = 0.6, and the case-control VE 0.32 and 0.39. As in GoGoVax, the two arms’ cumulative incidence of first diagnosis rose close together throughout follow-up (Figure S6). The result depended on intense exposure in the trial group: light encounters carrying 14% of its NG lowered *P*^-^*_e_* to 0.76 and raised the trial VE to 0.17 and 0.21, and with light encounters carrying 72% (*P*^-^*_e_* 0.28), or in the equal-intensity worlds (*P*^-^*_e_* 0.55), it was 0.30–0.41, above GoGoVax’s 95% CI. Activity weights made little difference (0.13 and 0.15).

In every world, an all-or-nothing vaccine without blunting was ruled out by the trials, since exposure intensity cannot lower its protection: its trial VE was 0.46 with *V*_0_ = 0.4 and 0.57 with *V*_0_ = 0.5.

Blunting by prior infection reconciled the trials with the observational studies under leaky protection, but all-or-nothing protection needed nearly every enrollee to be blunted. Full blunting over the 12 months before dose 1, which blunted 69% of trial enrollees, brought a leaky vaccine’s trial VE to 0.05 and 0.07, with the case-control VE at 0.32 and 0.39. All-or-nothing protection agreed only when blunting covered GoGoVax’s whole 18-month eligibility window, which blunted 88% of enrollees (0.02 and 0.06). A leaky vaccine needed less blunting, because heterogeneity in exposure intensity already lowered its trial VE: with *V*_0_ = 0.5, full blunting over 3–6 months or half blunting over 6–12 months gave a trial VE of 0.06–0.10, within GoGoVax’s 68% CI (Figure S3). Full blunting by NG at vaccination only, which affected 22% of enrollees, gave 0.10, within GoGoVax’s 68% CI, with the case-control VE at 0.30; with *V*_0_ = 0.6 it gave 0.14, at GoGoVax’s edge. Long blunting did not need heterogeneity in exposure intensity: in the equal-intensity worlds, full blunting over 12 months gave a trial VE of 0.08 and 0.10, within GoGoVax’s 68% CI, but blunting at a dose gave 0.25 and 0.32. Short blunting agreed only together with heterogeneity in exposure intensity.

Figures 3 and 4 show simulations of the England-like programme for one vaccine model per hypothesis: in the differential-intensity worlds, leaky protection without blunting and with full blunting at a dose or over 12 months, and in the equal-intensity worlds, blunting over 12 months.

**Figure 3:**
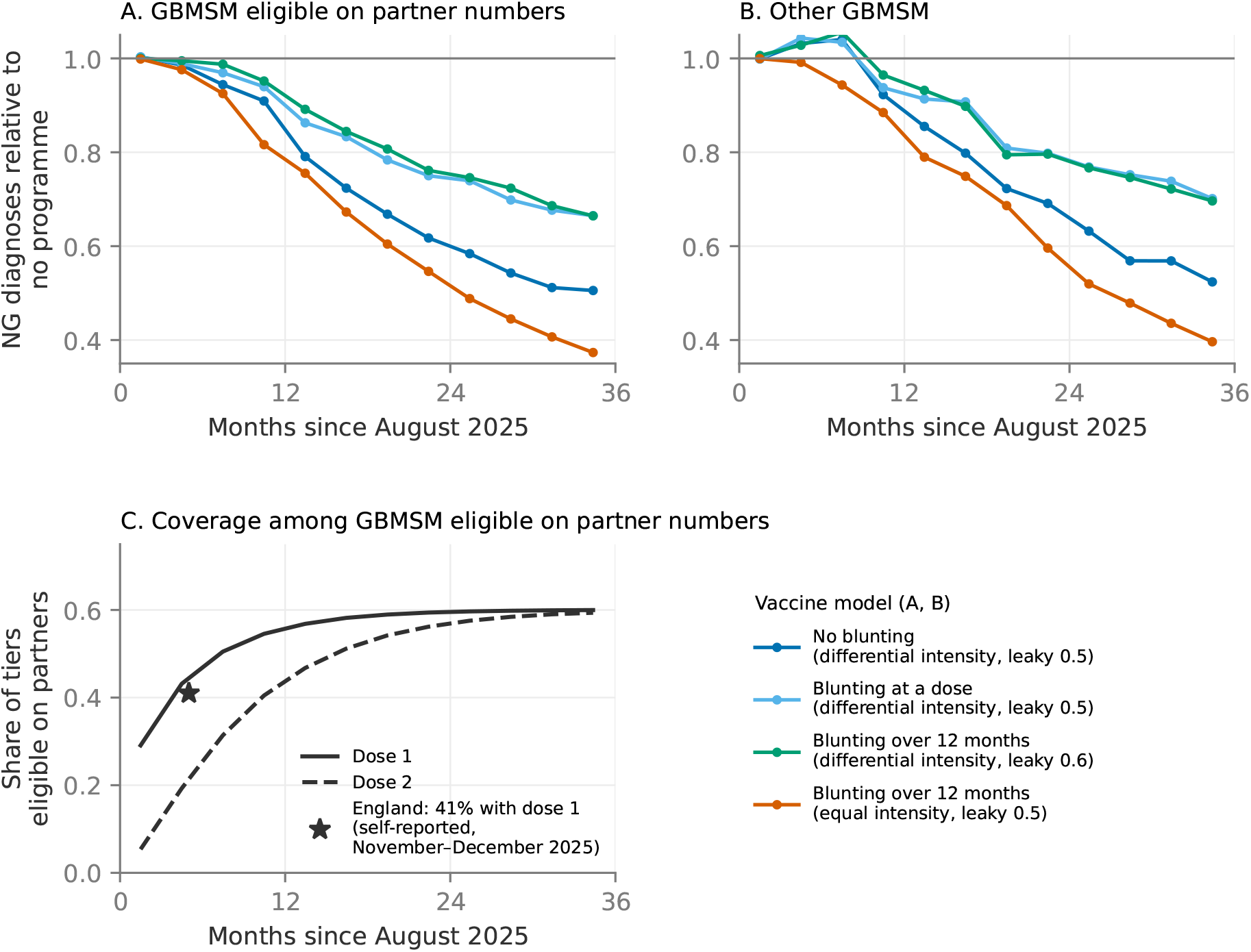
Impact of a programme like England’s. A programme, started in August 2025 and run for three years, in a selection of the vaccine models that agree with the trials and the observational studies or come close. Each line pools the runs of one world and vaccine model, compared with corresponding runs without the programme that start from the same population and epidemic. Table S4 gives further results for runs with waning and with activity weights. (A, B) NG diagnoses relative to no programme among GBMSM eligible for the programme (A; the two highest tiers) and other GBMSM (B; the rest of the population). (C) Coverage with dose 1 and dose 2 among GBMSM eligible on partner numbers, the same in every world; the star marks England’s self-reported uptake of at least one dose, 41% in November–December 2025.^24^

**Figure 4:**
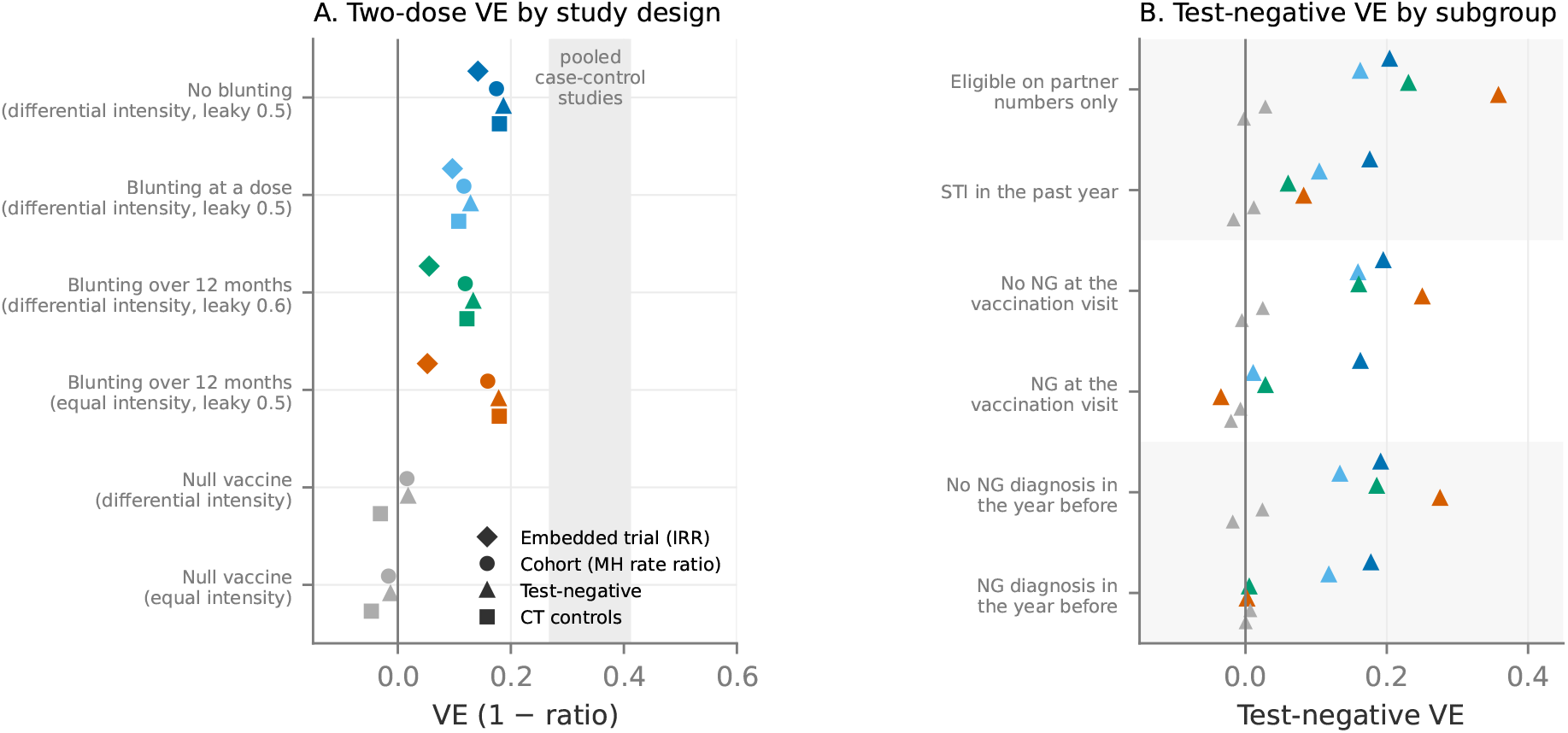
Estimated VE in a programme like England’s, for the vaccine models of Figure 3, in the same colours. Each marker pools the runs of one world and vaccine model. Grey markers show a null vaccine. (A) Two-dose VE as estimated from the embedded GoGoVax-sized trial and the three designs UKHSA plans: a cohort with vaccination as a time-varying exposure, a test-negative design, and a case-control design with CT cases as controls. The grey band shows the pooled real-world case-control estimate’s 95% CI, 0.27–0.41.^9^ (B) Test-negative VE within strata of vaccinees: by route of eligibility, by NG at the vaccination visit, and by an NG diagnosis in the year before. See the text for details.

In year 3, relative to the runs without the programme, NG diagnoses among GBMSM eligible on partner numbers fell by 30–46% in the differential-intensity worlds and by 57% in the equal-intensity worlds (Figure 3A). Blunting reduced the impact (30–31%, against 46% without), as did activity weights (15–33%) and waning (23–43%; Table S4). Diagnoses among other GBMSM, of whom 2–3% had two doses, fell almost as much (Figure 3B), so most of the effect was indirect: the fall was 4–6 times what two-dose coverage times the direct VE would give (about 3 times with activity weights).

The indirect effect also affected UKHSA’s two impact estimators^10^ (Table S4). In the differential-and equal-intensity worlds, comparing the change in diagnoses among targeted GBMSM with that among other GBMSM gave a ratio of 0.98–1.02 without waning, against 1.06–1.08 with a null vaccine, despite a true reduction of 30–57%, and VE back-calculated from the reduction and two-dose coverage read 0.50–0.97, against a direct VE of 0.1–0.2.

UKHSA’s three VE designs gave a two-dose VE of 0.10–0.19 for every vaccine model ( 0.05 to 0.02 with a null vaccine), at or above the embedded trial’s 0.05–0.14, because 40–48% of vaccinees qualified on partner numbers without a recent infection, whom the trial does not enrol (Figure 4A). This includes the observational studies’ own design, a case-control with CT cases as controls, which read 0.11–0.18 here against a pooled 0.34 (0.27–0.41) in the observational studies.

Without blunting, test-negative VE was the same in every stratum (0.16–0.20; Figure 4B). Both kinds of blunting lowered it among those with NG at the vaccination visit, to about zero ( 0.03 to 0.03, against 0.16–0.25 otherwise), and among those eligible through an STI (0.06–0.10, against 0.16–0.36). Only blunting over a window lowered it among those with an NG diagnosis in the year before (0.00–0.01, against 0.19–0.28; 0.12 and 0.13 with blunting at a dose). With every GBMSM diagnosis at England’s sexual health services, two years of data would detect each difference of 0.1 or more (at the 5% level) with a probability of at least 0.9 (at least 0.6 at half that scale; Supplement). With a null vaccine, the differences were at most 0.03.

## 4 Discussion

Using a transmission-modelling approach, we have shown that two biological mechanisms, namely vaccine blunting and heterogeneity in exposure intensity, can explain the apparent discrepancy in 4CMenB VE against NG between real-world RCT and observational estimates without the need to invoke confounding in the observational studies. If the RCT population’s exposure episodes were more likely to infect than the observational population’s, this alone brings a leaky vaccine’s simulated trial estimate to the upper edge of GoGoVax’s result. Long-lasting blunting by prior infection can reconcile RCT and observational estimates on its own. Together with heterogeneity in exposure intensity, blunting only by active NG at the time of vaccination was sufficient.

How long blunting lasts, if it exists, is the open question, and the evidence is sparse. MenGO’s hSBA data point to blunting only by active NG at vaccination,^12^ but this was based on a small sample of individuals (seven vaccinees with NG at vaccination), and it is unclear whether hSBA is a correlate of protection for NG.

In either of these biological mechanisms, the low VE found in the trial reflects the average protection that these individuals actually get. With a blunting mechanism, the average consists of some (blunted) individuals with little or no protection, and others with more; while with heterogeneity in exposure intensity, the effect is lower for everyone who is intensely exposed. Under either mechanism, the protection in the general population would conversely be higher, and of the magnitude measured by the real-world observational studies.

In simulations introducing a vaccine programme like England’s, NG diagnoses fell substantially, and among vaccinated and unvaccinated individuals in almost equal proportions. This held under heterogeneity in exposure intensity, blunting or both. Hence, even if the vaccine has no effect in a large share of vaccinees (blunting) or is greatly reduced among most or all of them (intense exposure), the overall effect is still large. This is an indirect effect: the most sexually active are net exporters of NG to the rest of the population. Even a small reduction in their transmission rate thus benefits everyone, and with the sequential nature of infection spread, small effects add up over time.

### 4.1 Limitations

We chose GoGoVax as the most stringent test of our hypotheses: it was the largest RCT and double-blind, and of the published RCTs it gave the lowest estimate with the narrowest confidence intervals. Pooling estimates of all four trials would have brought more of the hypotheses into agreement.

The differential-intensity worlds may overstate the heterogeneity in exposure intensity. Their casual encounters with an infectious partner infected far more often than NG positivity among MSM reporting sex with someone with NG would suggest.^25^ Simulated enrollees had as many partners as PROUD participants reported for condomless anal sex, but fewer than their partners of any kind,^26^ and adding low-risk encounters to make up the difference raised the simulated trial’s VE. In the observational group, surveys suggest more acts in persistent partnerships than we assumed,^27,28^ which would make its exposure episodes more likely to infect. Conversely, giving every activity tier of the trial group the same average risk per episode could hide heterogeneity within it that favours this explanation.

The model is also simpler than other current models, lacking two sexes, anatomical sites and population turnover,^22^ and has no partner notification, which acts only after infection. Protection did not wane in the simulated studies; observational data suggest waning only after about 2 years,^4,29^ beyond the trials’ follow-up, so waning would widen the discrepancy rather than explain it.

Beyond the designs we simulated, we did not consider whether differences in the real studies’ estimands could explain part of the discrepancy.

### 4.2 Conclusions

Our work shows that vaccine blunting and heterogeneity in exposure intensity are feasible explanations for the observed discrepancies between RCT and observational estimates of 4CMenB’s protection against NG. This should not be interpreted as proof that the explanations are correct. If true, a programme like England’s could reduce NG substantially even if VE estimated from programme data is low. Upcoming results from England and from the MAGI^30^ (completed, results not yet reported) and BIYELA^31^ (ongoing) trials, which enrol other populations, will hopefully shed more light on the problem. Heterogeneity in exposure intensity predicts a higher VE in populations with sexual behaviour associated with less intense exposure to NG, and blunting a higher VE where fewer enrollees were recently infected.

## Data Availability

The complete code used to produce the findings reported in this article can be found at https://github.com/folkehelseinstituttet/gonorrhoea_4CMenB_discrepancy_model. The study was solely model-based and did not involve any gathering of new data.

## Author contributions

JEM: Conceptualisation, Data curation, Formal analysis, Investigation, Methodology, Project administration, Resources, Software, Visualisation, Writing – original draft. GR: Conceptualisation, Methodology, Writing – review and editing. OB: Methodology, Writing – review and editing. VE: Methodology, Writing – review and editing. JMG: Methodology, Writing – review and editing. ML: Conceptualisation, Methodology, Project administration, Writing – original draft.

## Ethics

The study used no individual-level data and did not require ethical approval.

## Use of AI

Claude Code (Opus 5.5; Anthropic) was used to write and run code (Methods) as well as to aid in making figures and tables. It was also employed to review the manuscript text for consistency and completeness, suggesting phrasing in some places. The authors are fully responsible for all results and for the manuscript as written.

## Supplementary appendix

## S1 Model

### S1.1 Overview

The transmission model was an agent-based model of sexually active individuals. Its structure follows a line of individual-based models of gonorrhoea:^22,32–34^ an SIS infection on a dynamic network of persistent and one-off partnerships with concurrency, a small core of high-activity tiers mixing with the rest through one assortativity parameter, transmission per act, and diagnosis by symptoms or screening, with NG and CT on the same network as in Kretzschmar and colleagues^32^ and Jenness and colleagues.^34^ It is deliberately simpler than current gonorrhoea models: it has no sexes or ages, two transmission routes stand in for anatomical sites, and the population is closed and self-sustaining in its transmission, with no turnover by ageing, migration, births or deaths. It differs from them mainly in what it measures. Gonorrhoea vaccine models have traditionally taken vaccine efficacy as an input and projected impact;^35^ here a trial and an observational study are simulated in the same epidemic and analysed with the estimators the real studies used, so that the gap between them is an output. We deliberately wanted to keep the model simple while still capturing the main features of the population, networks and transmission dynamics relevant to the trial and observational settings. The model is thus to be viewed as a mechanism model with non-specific parameters, and the focus of the results is on the plausibility of different mechanisms, not on quantified predictions.

The main features and assumptions of the model are tabulated in Table S1. Figure S1 gives an overview of the model structure.

### S1.2 Populations and activity tiers

The simulations comprised two groups of 100,000 sexually active individuals each, which we refer to in the following as the “trial” and “observational” groups, respectively. The observational group was intended to represent a majority-heterosexual population of adolescents and young adults akin to the observational cohorts, while the trial group was intended to represent a GBMSM population, with the two highest tiers (15%) standing in for those on pre-exposure prophylaxis (PrEP) or living with HIV, from which the trial enrolled.

The sexual networks in the model were defined by agents being assigned to one of four activity tiers characterised by the rate of sexual acts of each agent, with 60%, 25%, 10% and 5% of the population in each tier ordered from least to most active. We number the tiers 0–3 in the observational group and 4–7 in the trial group, from least to most active. Act rates in each tier, before the group’s calibrated scale (below), were 0.0066, 0.026, 0.079 and 0.165 acts per person-day, respectively. Sexual acts were split between persistent partners, met repeatedly over months, and one-off casual encounters, possibly involving several acts. The groups differed in the share of acts in persistent partnerships, the number of acts per casual encounter, the per-act transmission probability and the symptomatic share of NG infections. In the trial group 30% of an agent’s acts were in persistent partnerships, against 70% in the observational group. Each group’s partnership formation rates, casual and persistent, were multiplied by a scale common to all its tiers and calibrated to the group’s NG incidence (below), so that partner numbers and acts scaled together while acts per partnership stayed fixed. The activity of each tier relative to the other tiers was thus the same in both groups.

### S1.3 Partnerships and mixing

The agents mixed assortatively in tiers by assigning 50% of partnership-network stubs (one end of a partnership still to be formed) to a within-tier pool where they were randomly paired each day, while the rest of the stubs were assigned to the group’s common pool, where they were paired at random, so partners came from each tier in proportion to its share of the pool’s stubs (its total activity). Persistent and casual stubs were pooled and paired separately. This led to about 63% of partnerships being within a tier. The groups mixed by letting 2% of stubs that leave their tier’s pool go to a population-wide pool, so that about 1% of each group’s partnerships were with the other group. A persistent partnership had a duration drawn from a geometric distribution with mean 180 days and a Poisson-distributed number of acts with mean 0.15 a day. Concurrent partnerships were possible. Casual encounters lasted for one day; the number of acts was drawn as 1 + Poisson(*µ_c_* 1), where *µ_c_* denotes the mean number of acts per casual encounter, which was set to 4 in the trial group and 1.5 in the observational group. An encounter between the two groups took the larger value. The high number of acts in the trial group was meant to stand in for several anatomical sites per encounter. Each tier’s partnership rates followed from its act rate *A_k_* and its group’s persistent share of acts *c*: (1 *c*)*A_k_/µ_c_* casual encounters a day, and on average *cA_k_/*0.15 persistent partnerships at a time, formed at that number divided by the mean duration of 180 days, both then multiplied by the group’s scale.

**Figure S1:**
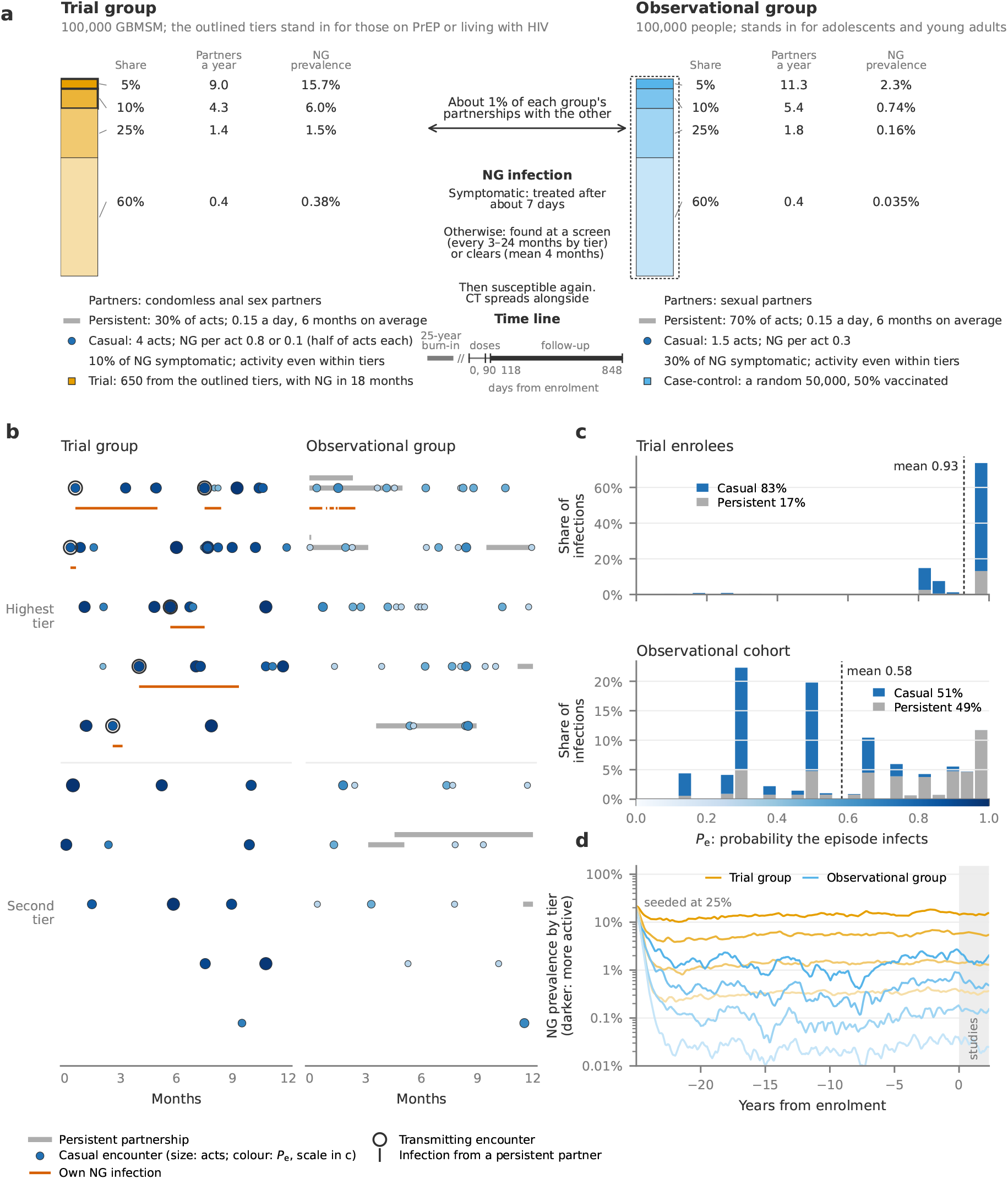
Model overview. The differential-intensity worlds, without a vaccine. Panels (a), (b) and (d) show one run in the observational setting with a per-act probability of 0.3 and 30% of NG symptomatic; (c) pools one run in each of the three plausible settings. (a) The two groups and their four activity tiers, with each tier’s partners a year and NG prevalence at enrolment. The trial enrolled from the trial group’s two highest tiers (outlined), which stand for GBMSM on PrEP or living with HIV; the case-control cohort (dotted outline) is a random half of the observational group. Centre: the course of an NG infection, mixing between the groups, and the study time line (doses on days 0 and 90; follow-up from day 118 to day 848). (b) One year of partnerships for ten people from each group’s two highest tiers: persistent partnerships (grey bars), casual encounters (circles, sized by number of acts and coloured by *p_e_*; scale in c) and NG infections (orange lines; outlined circles and ticks mark infections from a casual or a persistent partner). (c) The distribution of *p_e_*over exposure episodes, each weighted by its *p_e_*, so that the bars are shares of infections and their mean (dotted line) is *P*^-^*_e_*; trial enrollees and the observational cohort, by partnership type. _1_(_9_d) NG prevalence by tier from seeding through the 25-year burn-in to the study period (shaded); darker lines are more active tiers.

**Table S1:** The model’s main assumptions against data. The trial group stands in for gay, bisexual and other men who have sex with men (GBMSM), the observational group for adolescents and young adults. Model ranges are over the three observational settings. The comparison shows how the model’s assumptions line up against data. A trial-group partner stands for a condomless anal sex partner, as counted in PROUD. CT per-act probabilities are set per group to CT:NG ratios: 0.77 at enrolment (DOXYVAC baseline^11^) and 4.5 CT per NG diagnosis (3.5–6 in three studies^4,5,29^). NG: gonorrhoea; CT: chlamydia; PY: person-years; *P_e_*: probability that one exposure episode infects an unvaccinated person.

|  | Model | Data | Agreement |
|---|---|---|---|
| Populations | 100 000 per group in four activity tiers (5, 10, 25, 60%); about 1% of each group's partnerships are with the other group. The trial group's top two tiers stand in for men on PrEP or living with HIV | About 15% of UK GBMSM are eligible for England's programme (Green Book<sup>23</sup>) | Assumed |
| Partners, trial group | Mean 1.5 a year; 28% with 2 or more. Enrolees: median 2 (IQR 1–3) in 3 months | 2 or more a year: Natsal-3 13%, EMIS 25% (Prah 2016<sup>36</sup>). PROUD: 2 (1–5) receptive, 2 (1–6) insertive (Dolling 2016<sup>26</sup>) | Between surveys; spread narrow |
| Partners, observational | Mean 1.8–3.3 a year | Natsal-3, ages 16–24: 1.5–1.9 (reference tables<sup>37</sup>) | High |
| Encounters | Trial group: 30% of acts in persistent partnerships; casual encounters of 4 acts at several sites. Observational: 70% persistent; 1.5 acts. Mean *P<sub>e</sub>* 0.92 against 0.57 | Young adults: about 90% of acts in ongoing partnerships (Mercer 2017; Althaus 2015<sup>27,28</sup>). *P<sub>e</sub>* not measured | Persistent share low |
| NG course | Symptomatic 10% (trial group) or 30–70%, treated in about 7 days; otherwise found at screening (every 3–24 months) or cleared (mean 4 months) | Untreated: pharynx median 16 weeks (Barbee 2021<sup>38</sup>); rectum about 11 months (Chow 2016<sup>39</sup>) | Rectum longer |
| Trial group level | Placebo NG 44.6 per 100 PY; 88% with NG in the 18 months before; 22% infected at a dose. NG prevalence 1.7–2.0% | GoGoVax: 47.8, 89.6%, 26% (Seib 2026<sup>14</sup>). Prevalence 5.7% in community samples, 3.3% in Australia and New Zealand (Davies 2025<sup>40</sup>) | Trial matches; prevalence low |
| Observational level | NG 0.4–0.6 per 100 PY; prevalence 0.10–0.25%; CT prevalence 0.9–2.0% | 0.52 per 100 PY (Bruxvoort 2023<sup>6</sup>). Natsal-3: NG below 0.1%, CT 2.3–3.1% at 16–24 (Sonnenberg 2013<sup>41</sup>) | Incidence matches; NG high, CT low |
| Vaccine | Leaky, all-or-nothing or in between, efficacy 0.4–0.6. Blunting by NG at a dose or in the 3–18 months before | Response blunted by NG at a dose (*n* = 7), not by NG in the 3 months before (MenGO; Thng 2026<sup>12</sup>) | Tension for long windows |

### S1.4 Transmission, exposure episodes and *P*^-^*_e_*

In the trial group, each act independently went by a high-transmission route, with infection probability *p* = 0.8 per act if one partner was infectious and the other susceptible, or, with equal probability, by a low one with *p* = 0.1, standing in for the mix of acts and anatomical sites. In the observational group, *p* was 0.3 or 0.15, depending on the setting (below). An act across groups used the receiver’s group’s probability.

We defined an *exposure episode* as the acts in one partnership while one partner was NG-infectious and the other, the receiver, susceptible: one casual encounter, or a stretch of a persistent partnership that ended when the source cleared the infection or the partnership ended. The episode’s infection probability *p_e_*followed from the cumulative hazard *λ_e_* = Σ_acts_ ln(1 *p*_act_) for an unvaccinated receiver, *p_e_* = 1 *e^−λe^*. For example, at *p* = 0.3 a casual encounter of one act gives *p_e_* = 0.30; a trial-group casual encounter of four acts gives *p_e_* = 0.91; and a persistent partner whose asymptomatic infection lasts four months (about 18 acts) gives *p_e_* 1.

Each group’s exposure intensity can be summarised by the incidence-weighted mean *p_e_*, which we denote *P*^-^*_e_*, the mean of *p_e_* over episodes with an unvaccinated susceptible receiver, each weighted by its own *p_e_*:

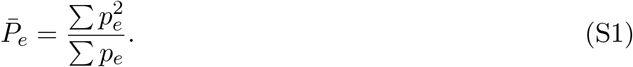

This definition lets *P*^-^*_e_* measure the mean infection probability *p_e_* of the episodes that produce cases. Since VE is estimated from cases, *P*^-^*_e_* is a useful measure to summarise the difference between the groups. In the differential-intensity worlds (Section S1.7), the trial group obtained a *P*^-^*_e_* of about 0.92, against 0.57 for the observational group. The higher *P*^-^*_e_* came mainly from the trial group’s casual encounters, with four acts on average at a mean per-act probability of 0.45, which carried 84% of its incidence; long asymptomatic infections in persistent partnerships added less.

### S1.5 Natural history, testing and simulation

The model simulated two co-circulating pathogens, *Neisseria gonorrhoeae* (NG) and *Chlamydia trachomatis* (CT). NG was the pathogen under study, while CT supplied the controls for the case-control study in the observational VE estimation (below). The natural history of disease for both pathogens was modelled as susceptible-infectious-susceptible (SIS). The duration of untreated NG infection was Gamma distributed with mean 120 days and a shape parameter of 2. Whether an NG infection was symptomatic depended on the infected person’s group: 10% were symptomatic in the trial group, against 30–70% in the observational group, depending on the setting (below). Symptomatic infection was treated after a Gamma-distributed period with mean 7 days and a shape parameter of 2. The duration of CT infection also followed a Gamma distribution, with mean 365 days and shape 2, 10% of cases symptomatic and treatment of symptomatic cases after a Gamma-distributed period of mean 10 days, shape 2. CT per-act probabilities were set per group to match CT:NG ratios: 0.77 at trial enrolment and 4.5 CT per NG diagnosis in the observational group (Table S1); in the verification runs they gave 0.73–0.81 and 3.4–8.5 across settings (Figure S2). Individuals were screened for infection at intervals with mean depending on tier: 24 months, 12 months, 6 months and 3 months for tiers ordered from least to most sexually active. For each agent, intervals were drawn from a geometric distribution with the given mean. The same interval applied to both groups. For individuals who were enrolled in a trial, screening was replaced by trial visits every 90 days. Every test, whether at screening, a trial visit or symptomatic presentation, was perfect: it detected and cleared all current NG and CT infections. There was no partner notification.

The model advanced in daily steps. Each run started with 25% of agents infected with NG and 25% with CT. An infection was infectious, and detectable, from the day after it was acquired. NG and CT were transmitted independently over the same acts; CT had one per-act probability per group, regardless of route. In all simulations we ran the model for a burn-in period of 25 years to reach equilibrium dynamics at the target incidence levels before starting the trial and observational study on day *T_e_*.

### S1.6 Observational settings and calibration

The details of heterosexual partnerships and acts are difficult to pin down from data, so we assumed four different settings for the model, defined by a per-act transmission probability of either *p* = 0.15 or *p* = 0.30, combined with a symptomatic share of NG infections of either 30% or 70%. Results were pooled over three of these settings, which we call the plausible settings; the combination *p* = 0.15 with 70% symptomatic was excluded because at calibrated incidence the observational group had 3.5 times as many partners a year as Natsal-3’s 16–24-year-olds (5.9, against 1.5 for women and 1.9 for men).^37,42^

Each group’s partner-rate scale was calibrated, jointly with its CT per-act probability (above), to an NG incidence of about 45 per 100 person-years (PY) of first diagnosis in the simulated trial’s placebo arm (close to GoGoVax’s 47.8) and 0.52 per 100 PY in the observational group (matching Bruxvoort and colleagues^6^). For each world we ran a grid of null-vaccine simulations over the two parameters, fitted smooth surfaces to log NG incidence and the log CT:NG ratio, took the values that met both targets and confirmed them in a verification run (Figure S2); a group that was the same in several worlds, such as the trial group of the differential-intensity worlds, was calibrated once.

The choice of 100,000 persons in each group was made for computational expedience; with two and four times the population, *P*^-^*_e_* and the trial group’s NG prevalence were unchanged, placebo incidence rose from 45 to 47 per 100 PY only because the trial became a smaller share of its tiers, and observational NG rose by about 25% in one setting.

### S1.7 Worlds and sensitivity analyses

We refer to each set of assumptions about the sexual networks, activity rates and transmission, in one observational setting, as a model *world*; worlds that differ only in their setting share a name, which is therefore plural. In our main analysis, the *differential-intensity worlds*, the trial group had a higher exposure intensity (*P*^-^*_e_* of 0.92 against 0.57 in the observational group). The alternative worlds where the groups share the same *P*^-^*_e_* (about 0.55) are referred to as *equal-intensity worlds*, and the worlds including light encounters as *light-encounter worlds*. Unless stated, results are for the three plausible settings, and the differential-intensity worlds are without activity weights or light encounters.

In alternative simulations, we stress-tested the assumptions of the model in various ways. In the equal-intensity worlds, we gave the trial group the observational group’s persistent share of acts, acts per casual encounter, per-act probability and symptomatic share, so that it differed only in partner numbers and had the same *P*^-^*_e_* as the observational group, about 0.55; because the trial group then took each setting’s observational parameters, it differed between settings.

In a further sensitivity analysis we added activity weights within tiers in the trial group of the differential-intensity worlds. Each person had a gamma-distributed weight *w* with mean 1 and shape *κ* and formed partnerships, persistent and casual, in proportion to *w*, so that partner numbers within a tier followed a negative binomial distribution with variance *µ* + *µ*^2^*/κ* while the tier’s mean rates were unchanged. We used *κ* = 0.5. Since the weights raise transmission, the trial group’s partner-rate scale was recalibrated to the same NG incidence and its CT per-act probability refitted.

In the light-encounter worlds, we included a third encounter type, “light encounters”, *e.g.* kissing and oral sex, with a per-encounter transmission probability *p_L_* of 0.02, 0.05 or 0.10, on top of activity weights, which raised the trial group’s partner numbers to those reported in DOXYVAC^11^ and PROUD.^26^ Light encounters were one-act encounters within the trial group, at a rate proportional to each tier’s casual-encounter rate, carrying no CT, with both groups’ scales refitted for each *p_L_*.

**Figure S2:**
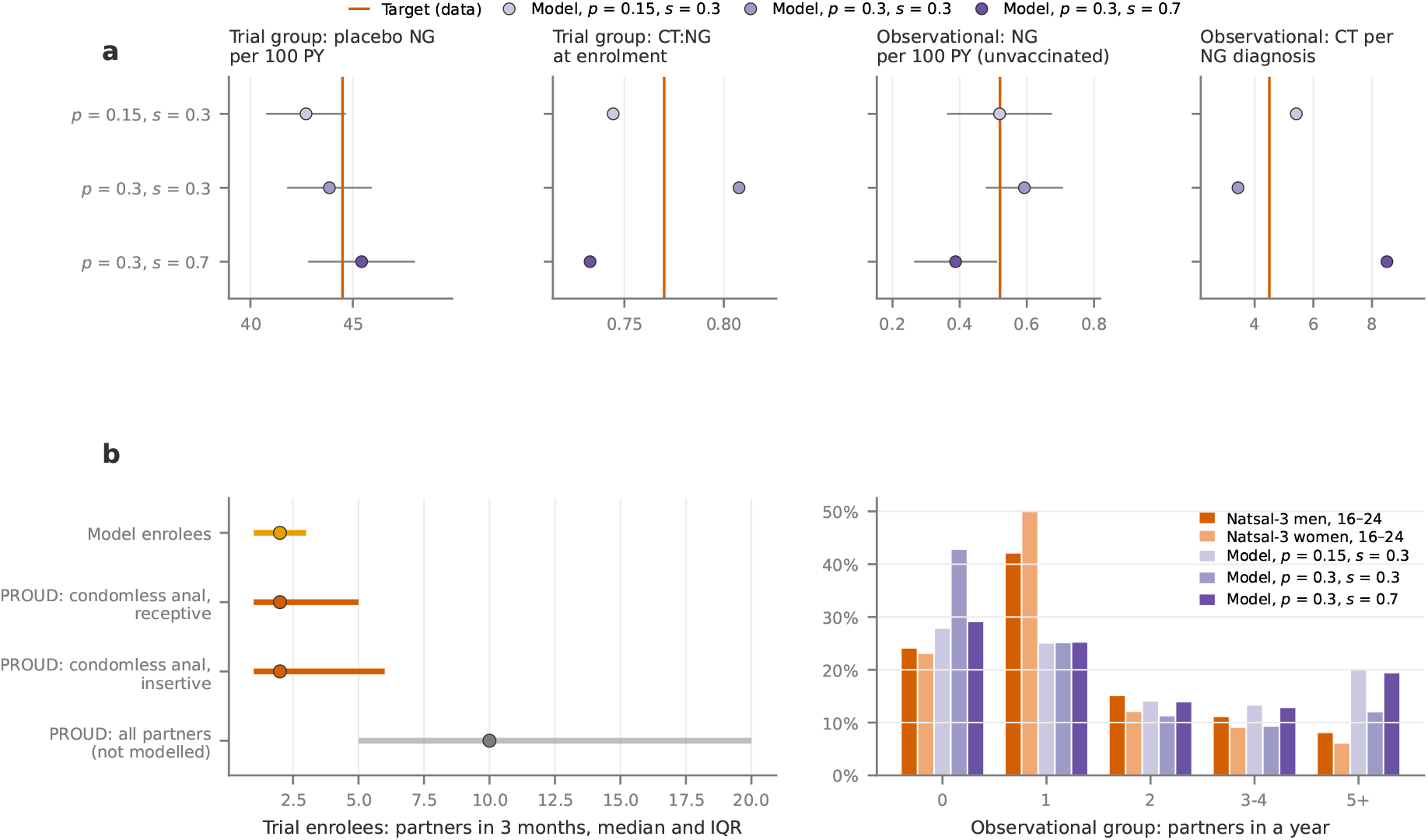
The differential-intensity worlds against their calibration targets and partner data. (a) The calibration targets (orange lines) in each plausible observational setting (*p*: per-act transmission probability; *s*: symptomatic share of NG in the observational group). In the trial group: NG diagnoses per 100 person-years (PY) in the simulated trial’s placebo arm (target 44.5; GoGoVax 47.8^14^), and the ratio of CT to NG prevalence among enrollees at enrolment (0.77). In the observational cohort: NG diagnoses per 100 PY among the unvaccinated (0.52^6^), and CT diagnoses per NG diagnosis (4.5; Table S1). Markers are means over 10 runs per setting without a vaccine, with 95% CIs of the mean; the CT ratios are pooled over the runs. (b) Partner numbers. Left: the trial enrollees’ persistent and casual partners in 3 months (median and interquartile range), against PROUD’s participants at baseline:^26^ condomless anal sex partners, receptive and insertive, which a trial-group partnership stands for, and all partners, which the model does not represent. Enrollees are pooled over the three settings, since the trial group is the same in each. Right: the share of the observational group with 0, 1, 2, 3–4 and 5 or more partners in a year, against men and women aged 16–24 in Natsal-3.^37^

## S2 Vaccine and hypotheses

### S2.1 Vaccine model

The vaccine acted only on acquisition: it did not change infectiousness, symptoms or the duration of infection. Each vaccinee had a protection level *v* that multiplied the per-act hazard by 1 *v*, so an exposure episode that would infect an unvaccinated receiver with probability *p_e_* infected the vaccinee with probability

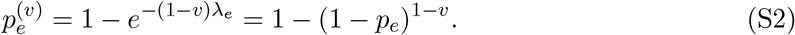

Scaling the hazard, rather than the per-act probability as in some individual-based models,^22^ matches compartmental models that scale the force of infection;^43^ under probability scaling the vaccine would protect more against acts with a high per-act probability. Protection levels were set by the vaccine parameter *V*_0_ and a mode parameter *α* [0, 1]. A share *V*_0_ of vaccinees (vaccine *takes*) had *v* = *V*_0_ + (1 *V*_0_)*α* and the rest (*failures*) *v* = (1 *α*)*V*_0_, so mean protection was *V*_0_ for every *α*. With *α* = 0 the vaccine is leaky, every vaccinee having *v* = *V*_0_; with *α* = 1 it is all-or-nothing (AON): takes are fully protected and failures not at all. We used *α* = 0, 0.5 and 1, with *V*_0_ = 0.5 or 0.6 for *α <* 1 and 0.4 or 0.5 for AON; only *α* = 0 and *α* = 1 are reported in the main manuscript.

Under a leaky vaccine, protection against an episode depends on how likely it was to infect:

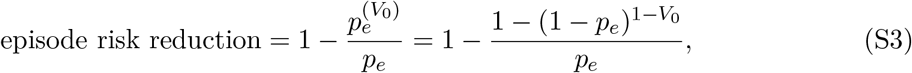

which is *V*_0_ as *p* 0 and falls to zero as *p* 1. At *V*_0_ = 0.5 it is 0.40 for an episode with *p_e_* = 0.57, the observational group’s *P*^-^*_e_*, and 0.22 for one with *p_e_* = 0.92, the trial group’s. Under AON action, the episode risk reduction is *V*_0_ whatever *p_e_*.

Vaccinees received two doses, on days *T_e_* and *T_e_* + 90, and were protected from 28 days after the second dose (day *T_e_* + 118), with no protection before. Protection did not wane, except in the simulations of an England-like programme (below). Most of follow-up fell within two years of vaccination, when South Australia’s programme found sustained protection against NG (VE 54% at 3–24 months, against 34% after 24 months),^29^ and the trial and the observational cohort shared the same follow-up window, so waning would have affected both alike.

### S2.2 Possible explanations

We investigated the following hypotheses to see if they can explain both the observational and RCT estimates.

#### Blunting

We assumed that the vaccine works less well in individuals who have had recent NG infection prior to vaccination. We modelled this using two parameters, the blunting effect and the blunting window. The blunting effect *b*, 0 *b* 1, is a factor multiplying the vaccine parameter *V*_0_ for vaccinees inside the blunting window – *b* = 0 means the vaccine is completely blunted and *b* = 1 means no blunting. The blunting window *T_b_* is the period before dose 1 in which an NG infection, diagnosed or not, blunts the response; in a variant, blunting is instead triggered by carrying NG at either dose. A blunted vaccinee had *V*_0_ replaced by *bV*_0_ before the protection level was drawn. For leaky vaccines we scanned *b* 0, 0.5, 1 with *T_b_* = 3, 6, 12 or 18 months or infection at a dose. For *α* = 0.5 and all-or-nothing vaccines we ran no blunting, full blunting (*b* = 0) over 3, 6, 12 or 18 months or at a dose, and half blunting (*b* = 0.5) over 6, 12 or 18 months, except that all-or-nothing protection with *V*_0_ = 0.4 was not run with full blunting at a dose or over 3 months. This gave 56 vaccine models in the differential-intensity worlds: 22 leaky, 18 intermediate and 16 all-or-nothing.

#### Leaky vaccine and heterogeneity in exposure intensity

When a vaccine is modelled with a leaky mode of action, the effective protection it offers to an individual depends on the infection probability of the exposures the vaccinee meets (Eq. S3).^21,44^ The RCTs were conducted among GBMSM who were selected based on several factors that indicate high infection risk, such as history of prior NG infection, syphilis or living with HIV. It is thus plausible that these individuals on average experience a higher *P*^-^*_e_*, that is, exposure episodes more likely to infect, than their counterparts in the observational studies, which were conducted in majority-heterosexual populations and, crucially, without such inclusion criteria. The equal-intensity worlds, in which the two groups share the same *P*^-^*_e_*, serve as a control without this difference in intensity, and activity weights and light encounters test whether the result of the differential-intensity worlds survives more realistic partner numbers in the trial group.

## S3 Simulated studies and VE estimation

### S3.1 Trial

Each simulation contained a sub-simulation of a double-blind placebo-controlled trial. We considered GoGoVax^14^ to be the main test of the hypothesis on the trial side, as it was, with its double-blinded design, arguably the strongest of the randomised trials, and at the same time the one that found the lowest VE point estimate. We therefore aligned our simulated trial to GoGoVax. To this end, at *T_e_*, the day burn-in ends, 650 agents were drawn at random from the two highest-activity tiers of the trial group and assigned 1:1 to vaccination or placebo. 585 (90%) were drawn among those with at least one diagnosed NG infection in the past 18 months, while the remaining 65 (10%) were drawn among people without such diagnosis. The latter is meant to stand in for GoGoVax’s enrolment route by syphilis diagnosis. Vaccination was administered in a two-dose regimen with dose 1 and 2 given at day *T_e_* and *T_e_* + 90, respectively. Per-protocol follow-up started on day *T_e_* + 118 (the day protection started, 28 days after dose 2) and lasted for 730 days. The outcome was the first positive test for NG. There was no loss to follow-up during the trial. Enrollees were tested at the 90-day visits and at symptomatic presentation, and every test detected and cleared all current NG and CT infections. VE in the trial was estimated from the first-episode incidence rate ratio between the vaccine and placebo arms,

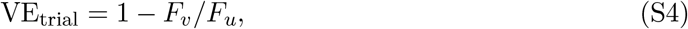

where *F_y_* = *C_y_/N_y_*, with *C_y_* the cumulative number of outcomes and *N_y_* the amount of person-time at risk, in vaccinated (*y* = *v*) and unvaccinated (*y* = *u*), respectively. This is in line with how the GoGoVax trial estimated VE. Model estimates were pooled over simulations by summing outcomes and person-time within each setting, and then averaged over the three settings.

### Simulated-trial protocol

#### Objective

To estimate, for each world and vaccine model, the per-protocol efficacy of two doses of 4CMenB against a first diagnosed NG infection that a trial built like GoGoVax^14^ would report.

#### Design

Individually randomised, placebo-controlled trial with 1:1 allocation. The trial runs inside a simulated population, one trial per simulation run. Allocation changes neither behaviour nor testing, so the trial is blinded by construction. Each vaccine model is run in the three plausible observational settings with 30 replicate trials each (independent seeds).

#### Setting

The trial runs in an agent-based model of NG and CT transmission. It has 200 000 people in two groups of 100 000, and about 99% of each group’s partnerships are within the group. The trial enrols from the trial group, which stands for gay, bisexual and other men who have sex with men (GBMSM); its two highest activity tiers stand for GBMSM on PrEP or living with HIV. Enrolment is on one day (day 0) after a 25-year burn-in to endemic equilibrium.

### Eligibility

- People in the trial group’s two highest activity tiers (15% of the group).
- At least one NG diagnosis in the 548 days (18 months) before day 0.
- 10% of enrolees are drawn from the same tiers without that diagnosis. This stands in for GoGoVax’s infectious-syphilis route.

#### Sample size

650 enrolees (325 per arm), a random sample of the roughly 5 000 eligible. This is 4.3% of the two tiers, small enough that the trial has little effect on transmission around it.

### Intervention

- *Vaccine arm:* two doses, on day 0 and day 90. Protection starts 28 days after dose 2 (day 118), stays constant to the end of follow-up, and is absent before then.
- *Placebo arm:* no protection.
- *Vaccine effect, set per vaccine model:*

**–** It acts on acquisition. Each vaccinee draws a protection *v* with mean *V*_0_, from leaky (every vaccinee *v* = *V*_0_, applied per act) to all-or-nothing (*v* = 1 with probability *V*_0_, otherwise 0).
**–** Under blunting, a vaccinee infected in a window of 3 to 18 months before dose 1, or, in a variant, infected at a vaccination visit, has mean protection *bV*_0_ instead of *V*_0_.

### Visits and testing

- Trial visits are every 90 days from day 0 to day 810 (10 visits, including both doses).
- Each visit tests for NG and CT. A test detects any current infection (perfect sensitivity, no anatomical sites), and treatment ends it the same day.
- Between visits, symptomatic infections present for testing after a delay.
- Routine testing is suspended while trial visits are scheduled and resumes after the last one.
- There is no loss to follow-up and there are no missed visits.

#### Outcome

The first NG infection diagnosed between day 119 and day 848, that is, days 1–730 after the per-protocol start (day 118). As in GoGoVax, an infection acquired before the per-protocol start but diagnosed after it counts.

#### Analysis population

All enrolees. Everyone receives both doses and completes follow-up, so the per-protocol and intention-to-treat populations coincide.

### Analysis

- *Estimate:* VE = 1 IRR. Each arm’s rate is its first diagnoses divided by its person-time, which runs from the per-protocol start to first diagnosis or day 730.
- *Per vaccine model:* events and person-time summed by arm over the 30 trials of each setting, and the resulting VEs averaged over the three settings. This estimates the expected trial result; its 95% CI comes from 500 bootstrap resamples of trials within each setting.
- *Between trials:* each trial’s VE; the spread of the 90 trials’ log rate ratios gives the agreement probabilities (below).

### S3.2 Case-control study

VE in the observational cohort was estimated in a case-control design as in three of the studies,^4,5,7^ with outcome defined as first positive test during the same 730-day window as the trial. Individuals testing positive for NG (regardless of CT coinfection at time of test) constituted cases, while individuals testing positive for CT and negative for NG constituted controls. Since the cited observational studies were heterogeneous in their design, cohort sizes, vaccination coverage and timing (Table S2), we needed to make some design choices for how to simulate an observational study in the model. Each agent in the 100,000-population observational group had a 50% chance of being part of the observed cohort, giving a cohort size of about 50,000 individuals (with reported VE estimates pooled over many simulations; below and Results). Each cohort member again had a 50% chance of being vaccinated; vaccination happened at the same schedule as the trial. This vaccinated about 25% of the group and lowered its NG incidence through an indirect effect; with 10% or 5% vaccinated, the case-control VE was on average 0.01–0.02 higher. The observational follow-up period was the same as in the trial. VE was estimated as one minus the odds ratio of being vaccinated in the case and control groups, respectively,

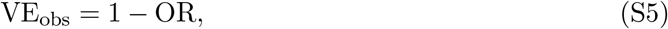

where the odds ratio was based on a 2 2 table of vaccination status by case or control for each simulation. Model estimates were pooled over simulations with the Mantel–Haenszel estimator, each simulation a stratum, over the runs of all three plausible settings.

#### Comparison with real-world studies

**Table S2:** Studies of meningococcal B vaccines against gonorrhoea (NG), and the simulated studies. VE is 1 minus the ratio given under estimand, as each paper reports it, with its 95% CI. PrEP: HIV pre-exposure prophylaxis. NAAT: nucleic acid amplification test. CT: chlamydia. MSM: men who have sex with men. OMV: outer membrane vesicle.

| Study; setting and population | Design; comparison | Blinding | Outcome | Estimand; estimator | VE (95% CI) | Size | Inclusion period | Stopping |
|---|---|---|---|---|---|---|---|---|
| *Randomised trials* |  |  |  |  |  |  |  |  |
| GoGoVax (Seib 2026);<sup>14</sup> Australia, multicentre. MSM with NG or infectious syphilis in the past 18 months, on PrEP or living with HIV | Individually randomised 1:1; 4CMenB, 2 doses (months 0 and 3), vs saline placebo | Double-blind (vaccinators unblinded, no role in outcome assessment) | First NG infection by NAAT (urogenital, anorectal or oropharyngeal) from 4 weeks after dose 2; screening every 3 months for 24 months, plus unscheduled visits | First-episode incidence rate ratio, per protocol (both doses, ≥2 follow-up visits); exact binomial CI; Poisson-adjusted as support | −0.5% (−26 to 20) | 654 randomised; 587 per protocol (296 vs 291); 315 first events (planned 730 and ≥251) | Jul 2021 – May 2023 | Fixed-event design; no interim efficacy analysis |
| DOXYVAC (Molina 2024);<sup>11</sup> Paris, 10 hospital sites. MSM on PrEP (ANRS PREVENIR cohort) with a bacterial STI in the past 12 months | 2 × 2 factorial; 4CMenB, 2 doses (months 0 and 2), vs no vaccine (1:1); doxycycline PEP vs none (2:1) | Open-label; laboratory and event review committee masked | First NG episode by PCR (throat, anus or urine) from month 3 (1 month after dose 2); visits every 3 months for 12–24 months | Hazard ratio, Cox adjusted for the other intervention; modified ITT (≥1 follow-up visit) | 22% (−1 to 40) | 556 randomised; 544 in the vaccine comparison (274 vs 270); 225 first events (planned 720) | Jan 2021 – Sep 2022 | Enrolment stopped early on the DSMB's advice after an unplanned interim analysis, which was later found to have omitted events |
| MenGO (Thng 2026);<sup>12</sup> Gold Coast, Australia, one centre. Gay and bisexual men aged 18–50 on PrEP or with NG in the past 3 months | Individually randomised 1:1; 4CMenB, 2 doses (months 0 and 3), vs no vaccine | Open-label; central laboratory | Primary: all NG episodes by NAAT from 30 days after dose 2 (sites at one visit count once). First episode also reported. Visits every 3 months for 2 years | Incidence rate ratio, Poisson (Andersen–Gill, robust SE), per protocol; first-episode IRR reported for comparison with DOXYVAC | All episodes, 22% (−51 to 60); first episode, 26% (−52 to 64) | 130 randomised (64 vs 66); 114 per protocol. Planned for VE 67% | Jan 2020 – Apr 2022 | DSMB oversight; none reported |
| Hong Kong (Kwan 2026);<sup>13</sup> one teaching hospital. MSM aged 18–50 at risk on PrEP-type criteria (STI history among them), NG-negative at recruitment, no earlier 4CMenB; 34% on PrEP, 7% living with HIV, 29% reporting an earlier NG diagnosis | Individually randomised 1:1 (blocks of 4); 4CMenB, 2 doses 1 month apart, vs saline placebo | Double-blind to month 12, then unblinded by design; 70% guessed their arm | First NG episode by NAAT (pharyngeal, rectal or urine) in the first year; no lag after dose 2 stated. Visits every 3 months for 2 years | Hazard ratio, Cox (age-adjusted as support); modified ITT (both doses, ≥1 follow-up test) | 24% (−87 to 69) | 161 given dose 1; 152 analysed (72 vs 80); 20 first events in year 1 (8 vs 12). Planned 150, for 1-year incidence 29% vs 10% | Screened May 2023 – Apr 2024 | Interim analysis by the DSMB when half had completed year 1; no stopping reported |
| *Observational studies* |  |  |  |  |  |  |  |  |
| Petousis-Harris 2017*;<sup>4</sup> New Zealand, 11 sexual-health clinics. Aged 15–30, born 1984–98, eligible for the 2004–06 MeNZB programme | Case-control, CT controls; MeNZB (an OMV vaccine), 3 doses ≥6 months before diagnosis, vs none | – | Case: laboratory-confirmed NG only (co-infection excluded); control: CT only. First recorded diagnosis per person | Odds ratio, logistic, adjusted for age, ethnicity, sex, region and deprivation | 31% (21 to 39) | 1 241 cases, 12 487 controls | Diagnoses 2004–2016 | – |
| Abara 2022*;<sup>5</sup> New York City and Philadelphia surveillance. Aged 16–23 | Case-control, CT controls, by infection; 4CMenB, 2 doses 30–180 days apart (or 1), vs none or before dose 1; infections ≥30 days after the last dose | – | Laboratory-confirmed NG vs CT; co-infection a separate outcome. Every infection, repeats included | Prevalence ratio of NG to CT, log-binomial GEE, adjusted for race and ethnicity, gender and city | 2 doses, 40% (23 to 53); 1 dose, 26% (12 to 37) | 18 099 NG, 124 876 CT and 24 731 co-infections in 109 737 people; 3 596 with 2 doses | Diagnoses 2016–2018 | – |
| Wang 2022*;<sup>7</sup> South Australia, school and young-adult programme from Feb 2019. Born 1998–2005 | Case-control, CT controls matched on date of birth (±28 days); 4CMenB, 2 doses (or ≥1), vs none; doses counted ≥6 months before onset | – | Case: notified NG (with or without CT); control: CT only. First episode | Odds ratio, logistic, adjusted | 33% (8 to 51) | 512 cases (575 episodes), 3 140 controls (3 847 episodes) | Notifications Feb 2019 – Jan 2021 | – |
| Raccagni 2023*;<sup>46</sup> Milan, one HIV clinic. MSM aged ≥18 living with HIV with an STI diagnosis in the period; STI testing every 6 or 12 months, or on demand | Unmatched case-control, controls with other STIs; 4CMenB, 2 doses (weeks 0 and 8), vs none. A vaccinated case's NG must follow dose 2; no lag, and no timing rule for controls, stated | – | Per person, over the whole period. Case: ≥1 NG by culture or NAAT (genital, rectal or pharyngeal), whatever other STIs; control: ≥1 syphilis, CT or anal HPV, and no NG | Odds ratio, logistic: unadjusted, and adjusted for age, years on antiretroviral therapy, CD4 and CD8 counts, HIV RNA and hepatitis co-infection; not for sexual behaviour | Adjusted, 44% (9 to 65); unadjusted, 42% (6 to 64) | 103 cases (24 vaccinated), 948 controls (325 vaccinated) | Diagnoses Jul 2016 – Feb 2021; median follow-up 3.8 years | – |
| Bruxvoort 2023;<sup>6</sup> Kaiser Permanente Southern California. Aged 15–30 | Matched cohort (1:4 on age, sex and year); ≥1 dose of 4CMenB vs MenACWY only (an active comparator) | – | First positive NG NAAT or culture ≥31 days after the index dose | Hazard ratio, Cox, adjusted; CT as negative-control outcome (HR 0.98) | 46% (14 to 66) | 6 641 vs 26 471; 27 vs 295 events | Vaccinated Jan 2016 – Dec 2019; follow-up to Aug 2020 | – |
| Robison 2023;<sup>8</sup> Oregon, two university outbreak campaigns that used both MenB vaccines. Vaccine recipients aged 18–29 | Cohort of vaccine recipients (the authors call it case-control); ≥1 dose of 4CMenB vs ≥1 dose of MenB-FHbp (a non-OMV active comparator); 57% and 53% had more than 1 dose | – | Reported NG from 1 month to 2 years after vaccination, or to study end | Cumulative incidence ratio, unadjusted (it reproduces the reported VE and CI from the counts); also for ages 18–19 alone. University tested as a confounder | 47% (13 to 68); ages 18–19, 59% (20 to 79) | 15 760 vs 15 212; 24 vs 44 cases | Campaigns from 2015 and 2016; follow-up to Mar 2018 | – |
| Abara 2024;<sup>47</sup> Kaiser Permanente Northern California. Aged 15–30 | Case-control, CT controls, by infection; 4CMenB, ≥1 dose, ≥14 days before, vs none | – | NG mono-infection vs CT mono-infection; co-infection separate. Repeats >30 days apart included | Prevalence ratio, log-binomial GEE; adjusted for race and ethnicity, or fully: also gender, insurance, deprivation and HIV | Adjusted for race and ethnicity, 23% (1 to 36); fully adjusted, 1% (−25 to 21) | 13 000 NG, 68 008 CT and 4 385 co-infections in 68 454 people; 558 vaccinated (0.8%) | Diagnoses 2016–2021 | – |
| Wang 2025;<sup>29</sup> South Australia, as Wang 2022, 5 years on. Born 1999–2009 | Case-control, CT controls matched on date of birth (±28 days, with replacement); 4CMenB, 2 doses, vs none; doses counted ≥3 months before onset | – | Case: first notified NG after vaccination (with or without CT); control: CT only | Odds ratio, logistic with SEs clustered by person, adjusted for age, sex and socioeconomic status | 39% (31 to 46) | 1 613 cases, 7 609 controls | Notifications Feb 2019 – Jan 2024 | – |
| McMillan 2026;<sup>48</sup> Northern Territory, Australia, with the Ngaanyatjarra Lands; school and community programme for ages 14–19, 2021–23. Aged 15–19 in the evaluation; 42% Aboriginal or Torres Strait Islander | Rolling cohort from linked notification and immunisation data; 4CMenB, after dose 1 and after dose 2, vs unvaccinated person-time | – | First positive NG test or notification from the start of at-risk time (1 Mar 2021 or the 15th birthday); no lag after a dose stated | Hazard ratio, Cox with time-varying vaccination, stratified by sex, age group and region; robust SEs. No data on sexual behaviour, STI history or testing | After dose 2, 38% (19 to 53); after dose 1, 27% (−3 to 48) | 30 650 people, 9.6% with 2 doses; 1 306 with NG | Mar 2021 – Feb 2024 | – |
| *Simulated studies (this paper)* |  |  |  |  |  |  |  |  |
| Simulated trial; the model's trial group (MSM on PrEP or living with HIV). Two highest activity tiers with an NG diagnosis in the past 18 months (10% without) | Individually randomised 1:1; 2 doses (days 0 and 90) vs placebo (no protection) | Blinded by construc-tion: allocation changes neither behaviour nor testing | First diagnosed NG episode from 28 days after dose 2 for 730 days; perfect tests every 90 days, plus symptomatic presentation | First-episode incidence rate ratio, all enrolees (per protocol = ITT). Events and person-time summed over 30 runs per setting, VE averaged over the 3 plausible settings; CI by bootstrap over runs | Expected VE, by assumptions (Figure S4): 0 to 57% over the 56 vaccine models of the differential-intensity worlds; 0 to 10% in the 18 that agree with both studies | 650 per run (325 vs 325); about 350 first events per run (280–390 by vaccine model and setting); 90 runs | One enrolment day, after a 25-year burn-in | None: fixed follow-up, no loss |
| Simulated case-control; the model's observational group (adolescents and young adults). A random half of the group (50 000 people) | Case-control, CT controls; 2 doses (days 0 and 90) to a random 50%, vs none | – | Each person's first NG or CT diagnosis in the trial's 730-day window. Case: NG (with or without CT); control: CT only | Odds ratio, unadjusted, Mantel–Haenszel over runs; CI by bootstrap over runs | Expected VE 30 to 46% over the 56 vaccine models; 30 to 40% in the 18 that agree with both studies | About 240 cases and 2 200 controls per run; 90 runs | One vaccination date; the same window as the trial | – |

\*Pooled in Wang 2024's random-effects meta-analysis<sup>9</sup> of case-control studies, fully vaccinated: VE 34% (27–41). South Australia entered through the programme's three-year analysis (Wang 2023<sup>45</sup>), not Wang 2022.

### S3.3 Pooling and confidence intervals

Each vaccine model was run with 30 seeds in each of the three plausible settings, and 95% CIs for the pooled estimates came from 500 bootstrap resamples of runs. These CIs show how well the runs determine the model’s expected estimate, which is far more precise than a single study.

### S3.4 Agreement criteria

Each vaccine model (mode of protection *α*, *V*_0_ and blunting) was judged against both designs at once. It agreed with the data if its case-control VE lay within the 95% CI of the pooled case-control estimate of a meta-analysis, 0.34 (0.27–0.41),^9^ and its trial VE within GoGoVax’s 68% CI ( 0.129 to 0.105);^14^ it was at the edge if the case-control VE was in range and the trial VE lay between GoGoVax’s 68% and 95% CIs (upper limit 0.20); and it disagreed otherwise. We used the 68% CI as the main criterion because a vaccine model whose expected trial VE lies near the 95% limit reproduces GoGoVax’s estimate only in an unusually low draw.

#### Agreement probabilities

We also assessed agreement in a second, continuous way that uses only the studies’ point estimates, GoGoVax’s VE of 0.5%^14^ and the pooled case-control VE of 0.34^9^ (Figure S5). Their sampling uncertainty is represented instead by the spread of the point estimate across simulated studies of the same precision. Simulated trials were already comparable to GoGoVax: the 90 trials of each vaccine model (30 runs in each of the three plausible observational settings) had a standard deviation of the log rate ratio of 0.09–0.13, against GoGoVax’s standard error of 0.12. A single simulated case-control study had a standard error 2.4–3.0 times the pooled estimate’s (0.13–0.16 against 0.054). We therefore pooled the case-control tables of *k* randomly drawn runs (Mantel–Haenszel; without replacement, across the three settings), with *k* (6–9 by vaccine model) chosen to match the pooled estimate’s precision, 4,000 times for each of the 56 vaccine models in the differential-intensity worlds. For the trials, we fitted a normal distribution to the 90 log rate ratios and took *z*_trial_ = (log 1.005 *m*)*/s*, with *m* and *s* its mean and standard deviation and log 1.005 GoGoVax’s log rate ratio; *z*_cc_ came likewise from the log odds ratios of the simulated meta-analyses and the pooled estimate’s, log 0.66. Each probability was two-sided, *p* = 2Φ(*z*); the corresponding shares of simulated estimates agreed with these to within 0.08 for the trial and 0.05 for the case-control study. The trial and the case-control study were drawn from different groups, and their estimates were uncorrelated across runs (mean correlation 0.03), so we combined them as *p_joint_ = P(X^2^_2_ ≥ z^2^_trial_ + z^2^_cc_)*: the probability that the vaccine model gives a trial and a pooled case-control estimate that together lie at least as far from its mean estimates, in standard deviations, as GoGoVax’s and the pooled estimate do. These probabilities quantify how surprising the real-world data would be under a vaccine model, not how likely the model is.

## S4 The England-like programme

### S4.1 Programme

England has offered 4CMenB to GBMSM since 1 August 2025.^10,23^ The programme is opportunistic and run at sexual health services, targeting GBMSM who have had a bacterial STI in the past 12 months or at least 5 sexual partners in 3 months (about 15% of UK GBMSM).^23^ Two doses are given at least 4 weeks apart, the first often at the attendance where an STI is tested for or treated.

We simulated such a programme in the trial group, excluding the trial’s enrollees, from day *T_e_*, which corresponds to 1 August 2025. The group’s two highest tiers, 15% of it, stood for those eligible on partner numbers. A person was eligible at a visit (a background test or a presentation with symptoms) if they were in these tiers or had an NG or CT diagnosis in the 365 days up to and including the visit, CT standing in for the other bacterial STIs. Each agent had a fixed acceptance trait, drawn independently of tier and infection history, so uptake was not confounded. Acceptors received dose 1 at their first eligible visit and dose 2 at their first visit at least 28 days later; the others were never vaccinated. The share of acceptors, 60%, was the only parameter set to match England’s data: it gave 39–40% of the top two tiers dose 1 by about month 5, against 41% self-reported in a community survey^24^ (Figure 3C). Second-dose uptake was 37–39% of first doses by then, against England’s 38%. The model gave a larger share of its first doses early: per month, its first doses in January–March 2026 were 0.40 of those in August–December 2025, against 0.55 in the doses England reported.^49,50^ Protection started 28 days after dose 2, one dose giving none, and followed the same vaccine model as in the trial simulations; the blunting window was counted back from dose 1, and blunting at a dose meant NG at dose 1 or dose 2. In some simulations protection waned: each vaccinee lost all of it after an exponentially distributed time with a mean of 3 years. The mean is an assumption; the modelling behind England’s programme assumed 31% protection lasting 18 months.^43^ DoxyPEP, demographic turnover and behaviour change were not modelled.

### S4.2 Simulations

The vaccine models were chosen to span the hypotheses (Section S3.4; Table S3). In the differential-intensity worlds they were leaky protection with *V*_0_ = 0.5 without blunting (at the edge); the same with full blunting at a dose (agrees); and leaky protection with *V*_0_ = 0.6 and full blunting over 12 months (agrees). In the equal-intensity worlds it was leaky protection with *V*_0_ = 0.5 and full blunting over 12 months. These four are shown in Figure 3. All but the 12-month row of the differential-intensity worlds were also run with waning, and the three differential-intensity rows without waning were rerun with activity weights (*κ* = 0.5), as a lower bound on impact (Table S4). Each run lasted three years, included a GoGoVax-sized trial enrolled on day *T_e_*, and was paired with a null-vaccine run on the same seed.

Every run used the calibrated world of its row (Section S1.6), with the population and burn-in of the trial simulations. The trial’s follow-up ended 848 days after *T_e_*, and the run continued to 1096 days after *T_e_*. Acceptance traits are shared within a pair, and so are all other random draws until protection first changes an infection; every later draw then differs. Paired runs thus start from the same population and epidemic but diverge after *T_e_*, and by year 3 they were no more alike than unpaired runs: the programme’s effect is estimated from runs pooled over seeds, not seed by seed. With waning, 72%, 51% and 37% of vaccinees kept their protection one, two and three years after it started. In the differential-intensity worlds one observational setting was run, because the trial group, and so the programme, is the same in every setting. In the equal-intensity worlds the trial group differs by setting, so the three plausible settings were run. Every test in the programme’s population was recorded from 18 months before *T_e_*. This allowed eligibility, the eligibility route and prior diagnoses to be reconstructed from the records, as a surveillance system would.

### S4.3 Outcomes

We report three outcomes (Figures 3 and 4):

- Impact (Figure 3A and B): NG diagnoses relative to the paired null runs, by quarter in the top two tiers and in the other two tiers of the trial group, and in the third year (Table S4).
- VE (Figure 4A): two-dose VE as estimated by the embedded trial and by the three designs in UKHSA’s evaluation plan:^10^ a cohort from each person’s first eligible visit, with vaccination as a time-varying exposure; a test-negative design; and a case-control design with CT cases as controls. All three were stratified by tier, calendar time and time since entry.
- VE within strata (Figure 4B): the test-negative VE by eligibility route, by NG at the entry visit and by an NG diagnosis in the year before. To estimate whether England’s data could detect these splits, we treated each run as a small England and scaled the run-to-run spread to England’s number of GBMSM diagnoses.^49,50^

The subsections below give details, and define the secondary measures we report in the text: UKHSA’s two impact estimators, which are the change in diagnoses in the top two tiers relative to that in the other two and VE back-calculated from impact and coverage; the fall in diagnoses relative to coverage times the cohort’s VE; the reproduction number of NG among GBMSM, from reruns with infection sources recorded (Table S5); and VE with doses counted as in UKHSA’s plan, from 14 days after each dose.

**Table S3:** Simulations of the England-like programme. Each vaccine row was paired with a null vaccine on the same seeds. Protection is leaky with the *V*_0_ shown, and *b* = 0 is full blunting. “Over 12 months” means blunting by NG infection in the 12 months before dose 1, and “at a dose” blunting by NG at dose 1 or dose 2. Waning is exponential with a mean of 3 years. *s*: symptomatic share in the observational group.

| World | Setting | Seeds per row | Vaccine rows |
|---|---|---|---|
| Differential intensity | *p* = 0.3, *s* = 0.3 | 16 | Leaky 0.5; leaky 0.5, *b* = 0 at a dose; leaky 0.6, *b* = 0 over 12 months; the first two with waning |
| Equal intensity | Three plausible | 11 per setting | Leaky 0.5, *b* = 0 over 12 months, with and without waning |
| Activity weights (*κ* = 0.5) | *p* = 0.3, *s* = 0.3 | 16 | Leaky 0.5; leaky 0.5, *b* = 0 at a dose; leaky 0.6, *b* = 0 over 12 months |

### S4.4 Impact

Impact was diagnosed NG in the third year (days 728–1091 after *T_e_*, four quarters of 91 days). It was summed over seeds and divided by the same sum in the paired null runs. Impact was reported for the top two tiers and the other tiers of the trial group. We also computed UKHSA’s two impact estimators^10^ and the amplification (Table S4).

- The ratio of changes: diagnoses in the top two tiers in year 3 over the year before *T_e_*, divided by the same ratio in the other tiers, averaged over runs. These tiers stand in for non-eligible GBMSM, although in the differential-intensity worlds 2.7% of them were vaccinated by the end of the run through the STI route.
- Back-calculated VE: one minus the impact, divided by the mean coverage with dose 2 in the top two tiers in year 3. We used the paired impact, not an extrapolated trend.

The amplification was the fall in diagnoses, one minus the impact, divided by the dose-2 coverage times the direct VE (the cohort design below), which is the same as the back-calculated VE divided by the direct VE. An amplification of 1 would mean that the fall is about what direct protection of the vaccinated alone gives.

**Table S4:**
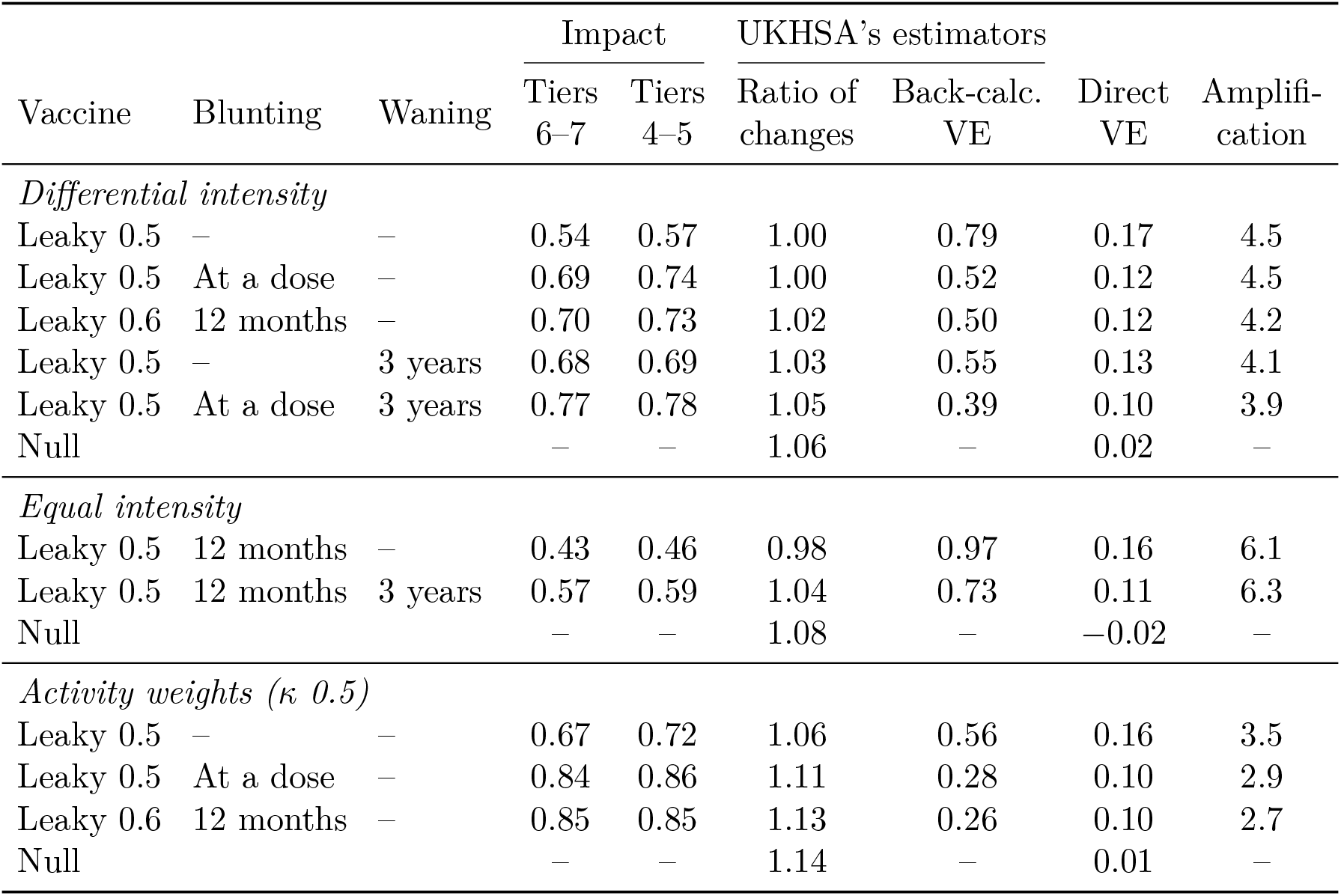
Impact measures of the England-like programme for the vaccine models of Figure 3, with their waning and activity-weights variants and each world’s null vaccine. Impact: NG diagnoses in year 3 relative to the paired runs without the programme, in tiers 6–7 (GBMSM eligible on partner numbers; Figure 3A) and tiers 4–5 (other GBMSM; Figure 3B). Ratio of changes: diagnoses in tiers 6–7 in year 3 over the year before the programme, divided by the same ratio in tiers 4–5, averaged over runs; 1 means the two groups changed alike. Back-calculated VE: one minus the impact in tiers 6–7, divided by their two-dose coverage in year 3 (58–59% in every row). Direct VE: the cohort design (Figure 4A). Amplification: the back-calculated VE divided by the direct VE. Blunting is full (*b* = 0); waning has a mean of 3 years. The equal-intensity rows are pooled over the three plausible observational settings.

|  |  |  | Impact |  | UKHSA’s estimators |  |  |  |
|---|---|---|---|---|---|---|---|---|
| Vaccine | Blunting | Waning | Tiers 6–7 | Tiers 4–5 | Ratio of changes | Back-calc. VE | Direct VE | Amplifi-cation |
| *Differential intensity* |  |  |  |  |  |  |  |  |
| Leaky 0.5 | – | – | 0.54 | 0.57 | 1.00 | 0.79 | 0.17 | 4.5 |
| Leaky 0.5 | At a dose | – | 0.69 | 0.74 | 1.00 | 0.52 | 0.12 | 4.5 |
| Leaky 0.6 | 12 months | – | 0.70 | 0.73 | 1.02 | 0.50 | 0.12 | 4.2 |
| Leaky 0.5 | – | 3 years | 0.68 | 0.69 | 1.03 | 0.55 | 0.13 | 4.1 |
| Leaky 0.5 | At a dose | 3 years | 0.77 | 0.78 | 1.05 | 0.39 | 0.10 | 3.9 |
| Null |  |  | – | – | 1.06 | – | 0.02 | – |
| *Equal intensity* |  |  |  |  |  |  |  |  |
| Leaky 0.5 | 12 months | – | 0.43 | 0.46 | 0.98 | 0.97 | 0.16 | 6.1 |
| Leaky 0.5 | 12 months | 3 years | 0.57 | 0.59 | 1.04 | 0.73 | 0.11 | 6.3 |
| Null |  |  | – | – | 1.08 | – | −0.02 | – |
| *Activity weights (κ 0.5)* |  |  |  |  |  |  |  |  |
| Leaky 0.5 | – | – | 0.67 | 0.72 | 1.06 | 0.56 | 0.16 | 3.5 |
| Leaky 0.5 | At a dose | – | 0.84 | 0.86 | 1.11 | 0.28 | 0.10 | 2.9 |
| Leaky 0.6 | 12 months | – | 0.85 | 0.85 | 1.13 | 0.26 | 0.10 | 2.7 |
| Null |  |  | – | – | 1.14 | – | 0.01 | – |

### S4.5 Reproduction number

To understand the mechanics of the programme’s impact in the model, we counted, for each NG infection, the infections its source caused while that infection lasted, counting a receiver reinfected by the same persistent partner each time. Pooling these offspring by the source’s and the receiver’s tier gave the realised reproduction number by tier and a next-generation matrix, whose largest eigenvalue over tiers 4–7 we report as the reproduction number of NG among GBMSM. Infections were grouped by when they were acquired (months 4–12, 12–24 and 24–30 of the programme), and each vaccinated run was compared with its paired null run (Table S5).

### S4.6 UKHSA’s VE designs

#### Cohort

The cohort included everyone in the programme’s population from their first eligible visit (a background test or a presentation with symptoms) on or after day *T_e_*. For acceptors this was the visit of dose 1. Each person was followed to the end of the run. Outcomes were NG-positive visits after entry. Unvaccinated person-time ran from entry to dose 1, and protected person-time from the start of protection (28 days after dose 2) to the end of the run, so time after waning counted as vaccinated, as it would in surveillance data. The time between dose 1 and protection, and the tests within it, were left out. VE was one minus the Mantel–Haenszel rate ratio, stratified by quarter, tier and time since entry (0–30, 30–60, 60–90, 90–180, 180–365, 365–730 and over 730 days).

#### Case-control designs

The test-negative design took NG-positive visits after entry as cases and NG-negative background tests as controls. A second design took CT-positive, NG-negative visits as controls. Both were stratified by 14-day period, tier and time since entry, and VE was one minus the Mantel–Haenszel odds ratio. For every design the Mantel–Haenszel sums were added across seeds and settings, so the seed and the setting were further strata.

#### Time since entry

Stratifying on time since entry is needed for two reasons. NG risk is highest just after a treated infection, because an untreated persistent partner can reinfect (particularly since the model does not include partner notification). And protected time cannot start until 56 days after entry. Without this stratum, the null vaccine read VE up to 0.07 in the recently infected strata.

#### Splits

Each design was also run within strata of three variables fixed at entry:

- the eligibility route: an STI in the past year if the person had an NG or CT diagnosis in the 365 days up to and including the entry visit, and partner numbers only otherwise;
- NG detected at the entry visit;
- an NG diagnosis in the 365 days before the entry visit.

#### UKHSA’s dose classification

In a sensitivity analysis, vaccination status followed UKHSA’s plan: none until 14 days after dose 1, then one dose until 14 days after dose 2, then two doses. Each level, and one and two doses pooled, was compared with no dose, with the same strata. VE for at least one dose was 0.06–0.15 after three years and 0.00–0.06 after one, lower than two-dose VE because one dose gives no protection in the model.

### S4.7 Precision at England’s scale

Each run was treated as a small England. England’s GBMSM had 9 129 NG diagnoses at sexual health services in October–December 2025 and 7 938 in January–March 2026.^50^ Dividing their sum by the run’s diagnoses in the programme’s population in its second and third quarters (days 91–272 after *T_e_*, roughly the same months) gave England’s size in runs, *m*. This was 6.7 in the differential-intensity worlds and 4.6 in the equal-intensity worlds. For each split, the contrast Δ was the pooled VE in the first stratum minus that in the second. Its standard error at England’s size was SE = SD*/*√*m*, where SD is the standard deviation of single-run contrasts.

**Table S5:**
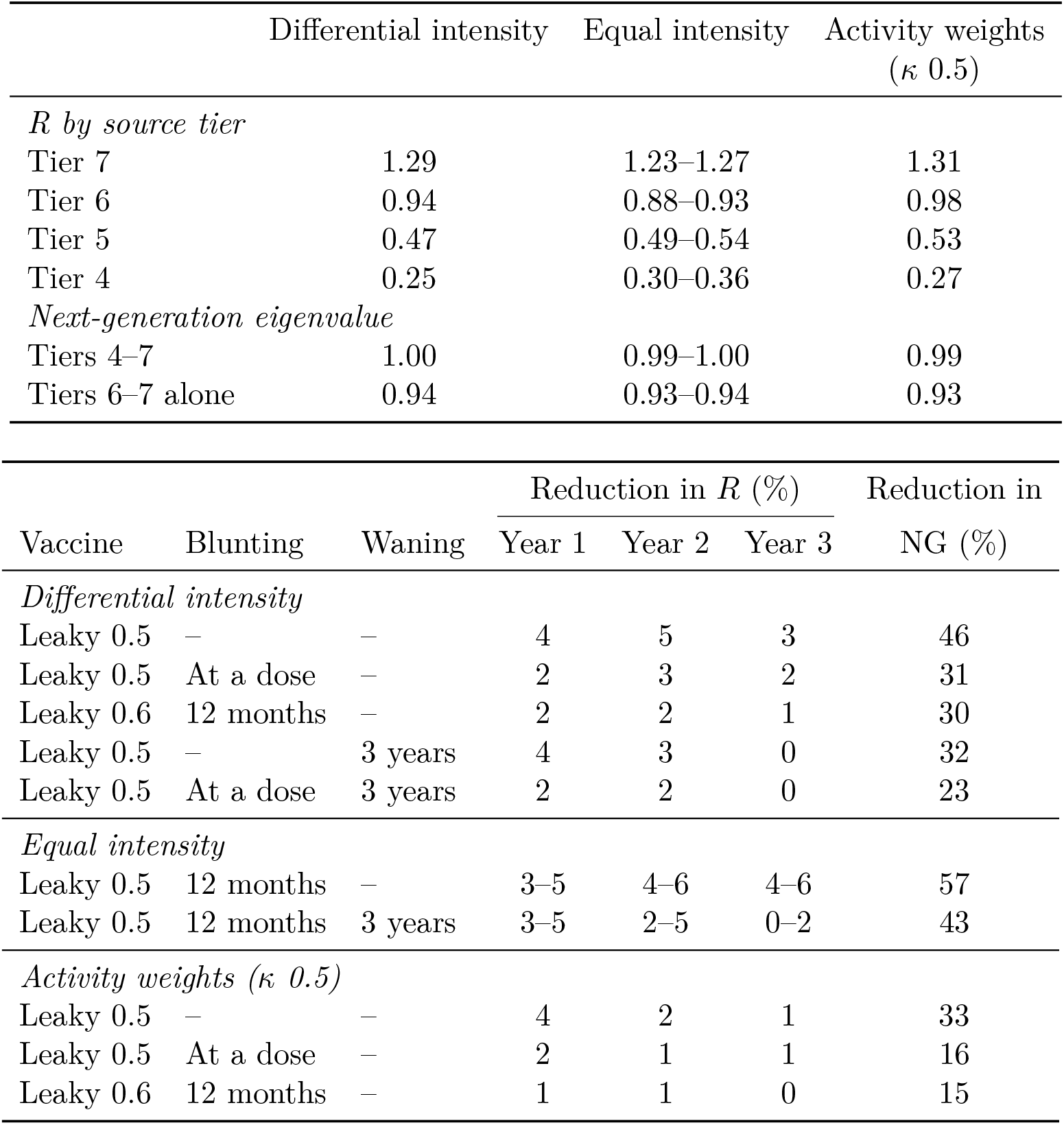
Reproduction number of NG in the England simulations. Top: without the programme, the realised reproduction number *R* by the source’s tier (the infections each NG infection causes while it lasts; tiers 6–7 are GBMSM eligible on partner numbers, tiers 4–5 other GBMSM), and the largest eigenvalue of the next-generation matrix over tiers 4–7 and over tiers 6–7 alone, for infections acquired in months 4–12. Bottom: the reduction in *R* of tiers 4–7 (the eigenvalue) relative to the paired runs without the programme, for infections acquired in months 4–12 (year 1), 12–24 (year 2) and 24–30 (year 3), and the fall in NG diagnoses in tiers 6–7 in year 3 relative to no programme (Figure 3A). Blunting is full (*b* = 0); waning has a mean of 3 years. Ranges are over the three observational settings of the equal-intensity worlds; reductions in *R* are precise to about one percentage point. Year 3 ends six months before the runs do, so that most of its infections have run their course.

|  | Differential intensity | Equal intensity | Activity weights (*κ* 0.5) |  |  |  |
|---|---|---|---|---|---|---|
| *R by source tier* |  |  |  |  |  |  |
| Tier 7 | 1.29 | 1.23–1.27 | 1.31 |  |  |  |
| Tier 6 | 0.94 | 0.88–0.93 | 0.98 |  |  |  |
| Tier 5 | 0.47 | 0.49–0.54 | 0.53 |  |  |  |
| Tier 4 | 0.25 | 0.30–0.36 | 0.27 |  |  |  |
| *Next-generation eigenvalue* |  |  |  |  |  |  |
| Tiers 4–7 | 1.00 | 0.99–1.00 | 0.99 |  |  |  |
| Tiers 6–7 alone | 0.94 | 0.93–0.94 | 0.93 |  |  |  |
|  |  |  | Reduction in *R* (%) |  |  | Reduction in NG (%) |
| Vaccine | Blunting | Waning | Year 1 | Year 2 | Year 3 |  |
| *Differential intensity* |  |  |  |  |  |  |
| Leaky 0.5 | – | – | 4 | 5 | 3 | 46 |
| Leaky 0.5 | At a dose | – | 2 | 3 | 2 | 31 |
| Leaky 0.6 | 12 months | – | 2 | 2 | 1 | 30 |
| Leaky 0.5 | – | 3 years | 4 | 3 | 0 | 32 |
| Leaky 0.5 | At a dose | 3 years | 2 | 2 | 0 | 23 |
| *Equal intensity* |  |  |  |  |  |  |
| Leaky 0.5 | 12 months | – | 3–5 | 4–6 | 4–6 | 57 |
| Leaky 0.5 | 12 months | 3 years | 3–5 | 2–5 | 0–2 | 43 |
| *Activity weights (κ 0.5)* |  |  |  |  |  |  |
| Leaky 0.5 | – | – | 4 | 2 | 1 | 33 |
| Leaky 0.5 | At a dose | – | 2 | 1 | 1 | 16 |
| Leaky 0.6 | 12 months | – | 1 | 1 | 0 | 15 |

In the equal-intensity worlds the SD was taken within settings, because their true VE differ. Power at the 5% level was Φ( Δ */*SE 1.96) + Φ( Δ */*SE 1.96). It was computed with one, two and three years of data. Counting every GBMSM diagnosis at a sexual health service as one in the modelled population is an upper bound. Each result was therefore repeated at half of England’s size (*m/*2), for linkage, missing data and GBMSM outside the modelled population. The SD comes from 16 or 33 runs and is uncertain by about 20%. Bias in England’s data, such as confounding of uptake, doxyPEP or differences in testing between vaccinees and others, is not included.

## S5 Literature search

We searched PubMed on 18 November 2026, without date or language restrictions, for publications on meningococcal group B vaccines and gonorrhoea, using the query below, and read titles and abstracts to find those relevant to our question. This was a targeted search, not a systematic review.

(“4CMenB”[tiab] OR Bexsero[tiab] OR MeNZB[tiab] OR “outer membrane vesicle”[tiab] OR “outer membrane vesicles”[tiab] OR “meningococcal B”[tiab] OR “meningococcal group B”[tiab] OR “serogroup B meningococcal”[tiab] OR MenB[tiab]) AND (gonorrh*[tiab] OR “Neisseria gonorrhoeae”[tiab] OR gonococc*[tiab]) AND vaccin*[tiab]

The search returned 158 results. Of these, 16 reported VE against gonorrhoea (four randomised trials and 12 observational publications from nine study populations), four were meta-analyses and eight were transmission models. Models of 4CMenB programmes have taken the observational VE as the vaccine’s protection, and we found no study that simulated a randomised trial and an observational study in the same model.

## S6 Supplementary results

Intermediate protection (*α* = 0.5) fell between leaky and all-or-nothing protection (Figure S4). Without blunting, its trial VE was 0.19 and 0.28 with *V*_0_ = 0.5 and 0.6. With full blunting over 12 months it fell to 0.06 and 0.08, within GoGoVax’s 68% CI, though with *V*_0_ = 0.6 the case-control VE, 0.41, lay just above the pooled CI; blunting at a dose was not enough (0.13 and 0.21).

**Figure S3:**
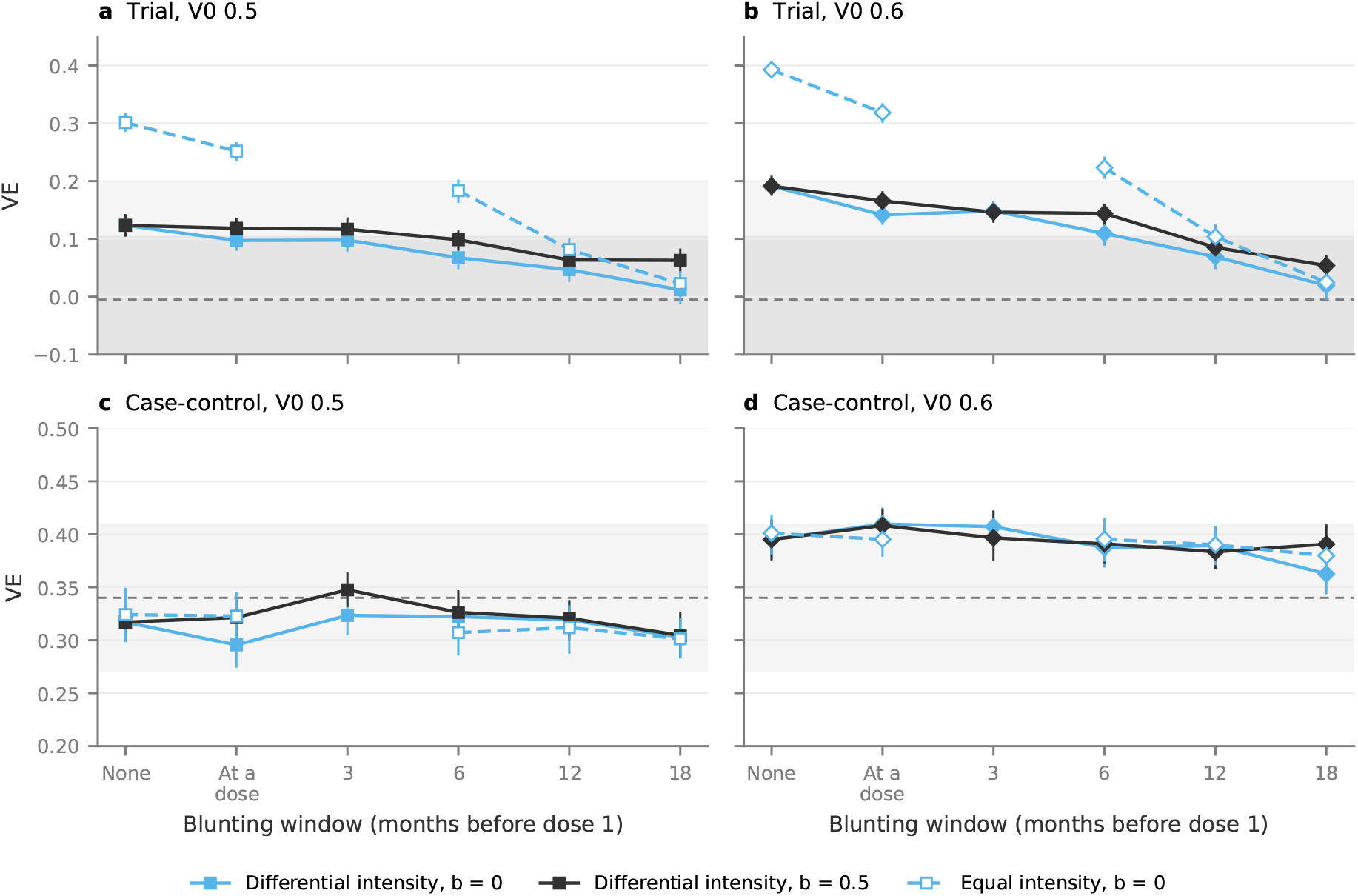
How long blunting must last. Estimated VE in the trial (a, b) and the observational case-control study (c, d) against the blunting window, for a leaky vaccine with *V*_0_ = 0.5 (a, c) and *V*_0_ = 0.6 (b, d). The window is the time before dose 1 in which an NG infection blunts the vaccine response; “at a dose” means blunting only by NG at dose 1 or dose 2, and “none” means no blunting, a point shared by the two differential-intensity lines. Lines show the differential-intensity worlds with full (*b* = 0) and half (*b* = 0.5) blunting, and the equal-intensity worlds with full blunting (dashed; not run at 3 months). Estimates are pooled over the three plausible observational settings, with 95% CIs from bootstrap resamples of runs. In (a) and (b), the dashed line and shaded bands show GoGoVax’s point estimate and its 68% and 95% CIs;^14^ in (c) and (d), the dashed line and shaded band show the pooled case-control estimate and its 95% CI, 0.34 (0.27–0.41).^9^

**Figure S4:**
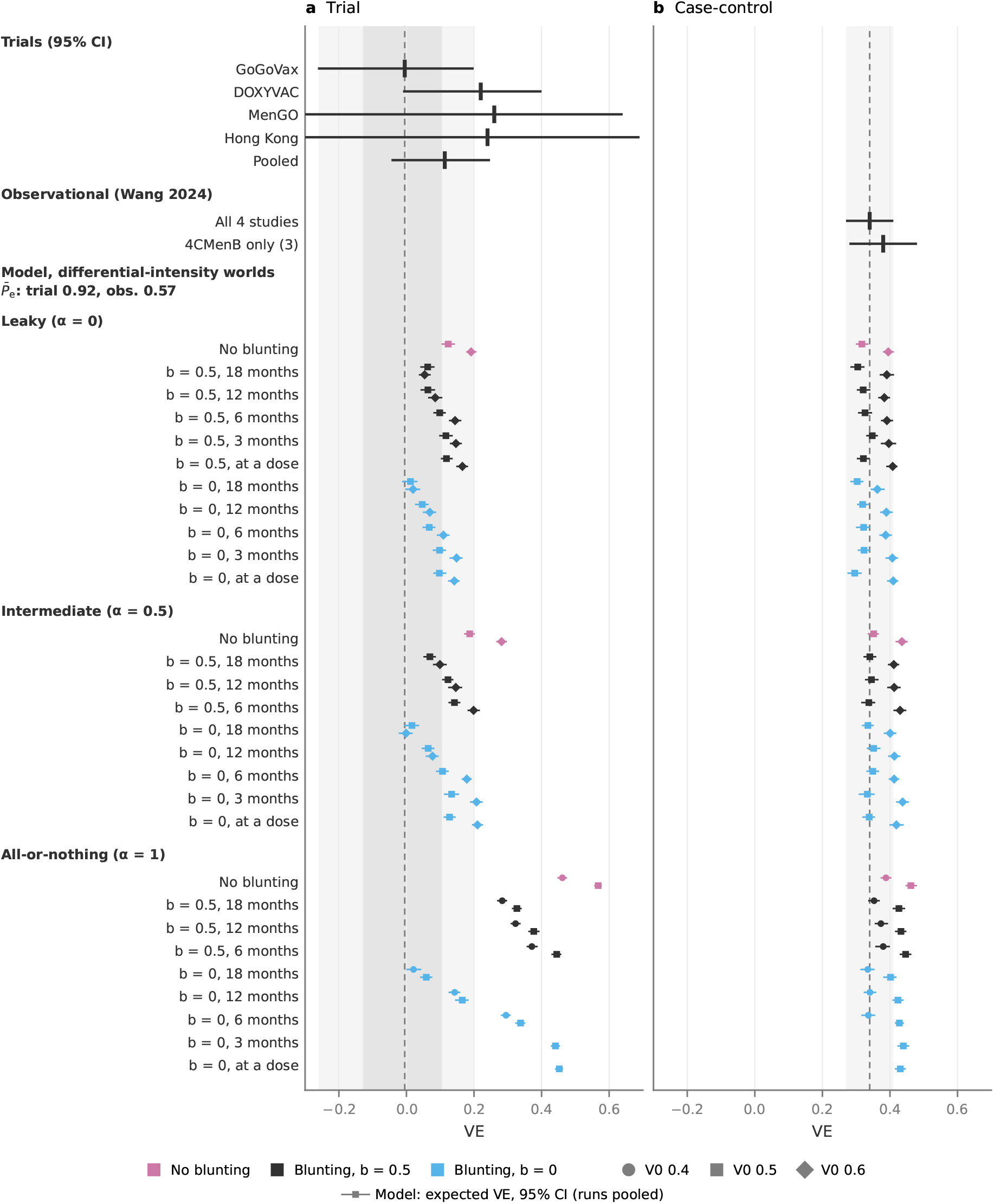
Estimated VE for every vaccine model in the differential-intensity worlds. (trial-group *P*^-^*_e_* 0.92, observational 0.57), in the trial (a) and the observational case-control study (b), pooled over the three plausible observational settings. Rows are grouped by mode of protection: leaky (*α* = 0), intermediate (*α* = 0.5) and all-or-nothing (*α* = 1). Each row is one blunting setting: none, half (*b* = 0.5) or full (*b* = 0), over the 18, 12, 6 or 3 months before dose 1 or by NG at a dose only. Marker shape gives *V*_0_ and colour the blunting. Markers, bars, shading and data as in Figure 2. Table S6 gives the estimates. all-or-nothing (*V*_0_ = 0.4) protection. All-or-nothing protection without blunting was incompatible with GoGoVax (*p <* 0.001).

The agreement probabilities (Section S3.4) largely matched the agreement classes (Figure S5 cf. Figure S4). All 18 vaccine models that agreed had a joint probability above 0.05, 12 of them above 0.32, and 20 of the 21 that disagreed had one below 0.05; the exception, intermediate protection with *V*_0_ = 0.6 and full blunting over 12 months (0.06), disagreed only because its case-control VE, 0.41, lay just above the pooled CI. Of the 17 at the edge, 7 had a joint probability above 0.32 and 4 below 0.05; the latter were all leaky with *V*_0_ = 0.6 and no or short blunting, and were moderately off on both sides. Under heterogeneity in exposure intensity alone (leaky protection, *V*_0_ = 0.5, no blunting), 10% of the simulated GoGoVax-sized trials found a VE as low as GoGoVax’s (*p*_trial_ = 0.18), while the pooled case-control estimate was typical of the model (*p*_cc_ = 0.51; joint 0.33). With full blunting over 12 months the joint probability rose to 0.72; it was highest, 0.97, with full blunting over 18 months under intermediate (*V*_0_ = 0.5) or

**Table S6:**
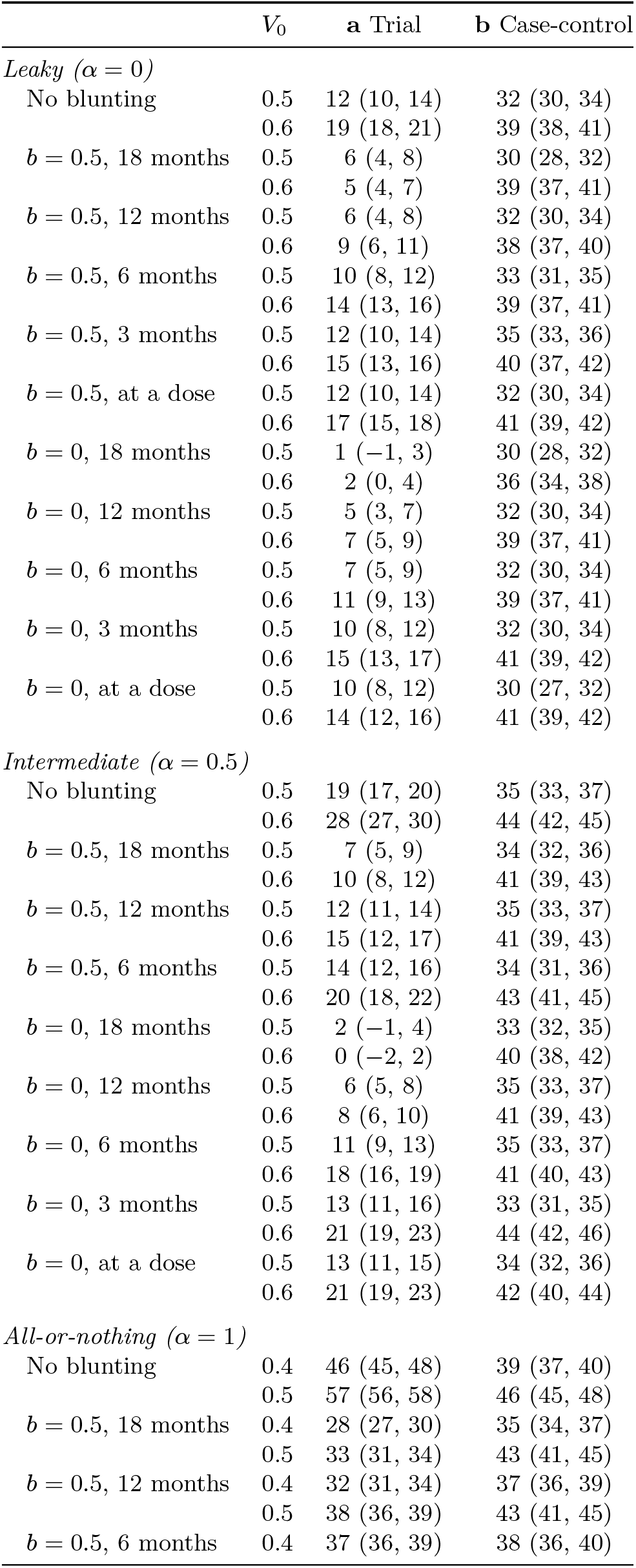
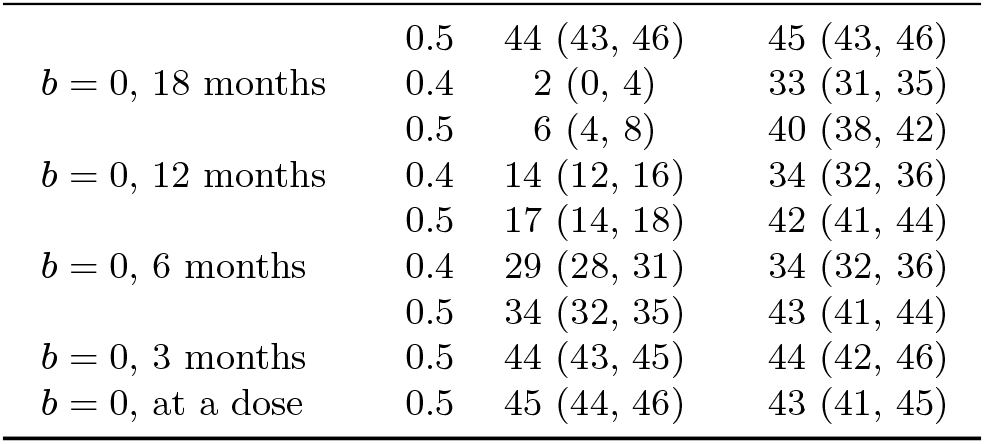
Estimated VE for every vaccine model in the differential-intensity worlds, in the trial (a) and the observational case-control study (b), as in Figure S4: VE in percent, with 95% CIs from bootstrap resamples of runs, pooled over the three plausible observational settings.

|  | *V*<sub>0</sub> | **a** Trial | **b** Case-control |
|---|---|---|---|
| *Leaky (α = 0)* |  |  |  |
| No blunting | 0.5 | 12 (10, 14) | 32 (30, 34) |
|  | 0.6 | 19 (18, 21) | 39 (38, 41) |
| ***b*** = 0.5, 18 months | 0.5 | 6 (4, 8) | 30 (28, 32) |
|  | 0.6 | 5 (4, 7) | 39 (37, 41) |
| ***b*** = 0.5, 12 months | 0.5 | 6 (4, 8) | 32 (30, 34) |
|  | 0.6 | 9 (6, 11) | 38 (37, 40) |
| ***b*** = 0.5, 6 months | 0.5 | 10 (8, 12) | 33 (31, 35) |
|  | 0.6 | 14 (13, 16) | 39 (37, 41) |
| ***b*** = 0.5, 3 months | 0.5 | 12 (10, 14) | 35 (33, 36) |
|  | 0.6 | 15 (13, 16) | 40 (37, 42) |
| ***b*** = 0.5, at a dose | 0.5 | 12 (10, 14) | 32 (30, 34) |
|  | 0.6 | 17 (15, 18) | 41 (39, 42) |
| ***b*** = 0, 18 months | 0.5 | 1 (−1, 3) | 30 (28, 32) |
|  | 0.6 | 2 (0, 4) | 36 (34, 38) |
| ***b*** = 0, 12 months | 0.5 | 5 (3, 7) | 32 (30, 34) |
|  | 0.6 | 7 (5, 9) | 39 (37, 41) |
| ***b*** = 0, 6 months | 0.5 | 7 (5, 9) | 32 (30, 34) |
|  | 0.6 | 11 (9, 13) | 39 (37, 41) |
| ***b*** = 0, 3 months | 0.5 | 10 (8, 12) | 32 (30, 34) |
|  | 0.6 | 15 (13, 17) | 41 (39, 42) |
| ***b*** = 0, at a dose | 0.5 | 10 (8, 12) | 30 (27, 32) |
|  | 0.6 | 14 (12, 16) | 41 (39, 42) |
| *Intermediate (α = 0.5)* |  |  |  |
| No blunting | 0.5 | 19 (17, 20) | 35 (33, 37) |
|  | 0.6 | 28 (27, 30) | 44 (42, 45) |
| ***b*** = 0.5, 18 months | 0.5 | 7 (5, 9) | 34 (32, 36) |
|  | 0.6 | 10 (8, 12) | 41 (39, 43) |
| ***b*** = 0.5, 12 months | 0.5 | 12 (11, 14) | 35 (33, 37) |
|  | 0.6 | 15 (12, 17) | 41 (39, 43) |
| ***b*** = 0.5, 6 months | 0.5 | 14 (12, 16) | 34 (31, 36) |
|  | 0.6 | 20 (18, 22) | 43 (41, 45) |
| ***b*** = 0, 18 months | 0.5 | 2 (−1, 4) | 33 (32, 35) |
|  | 0.6 | 0 (−2, 2) | 40 (38, 42) |
| ***b*** = 0, 12 months | 0.5 | 6 (5, 8) | 35 (33, 37) |
|  | 0.6 | 8 (6, 10) | 41 (39, 43) |
| ***b*** = 0, 6 months | 0.5 | 11 (9, 13) | 35 (33, 37) |
|  | 0.6 | 18 (16, 19) | 41 (40, 43) |
| ***b*** = 0, 3 months | 0.5 | 13 (11, 16) | 33 (31, 35) |
|  | 0.6 | 21 (19, 23) | 44 (42, 46) |
| ***b*** = 0, at a dose | 0.5 | 13 (11, 15) | 34 (32, 36) |
|  | 0.6 | 21 (19, 23) | 42 (40, 44) |
| *All-or-nothing (α = 1)* |  |  |  |
| No blunting | 0.4 | 46 (45, 48) | 39 (37, 40) |
|  | 0.5 | 57 (56, 58) | 46 (45, 48) |
| ***b*** = 0.5, 18 months | 0.4 | 28 (27, 30) | 35 (34, 37) |
|  | 0.5 | 33 (31, 34) | 43 (41, 45) |
| ***b*** = 0.5, 12 months | 0.4 | 32 (31, 34) | 37 (36, 39) |
|  | 0.5 | 38 (36, 39) | 43 (41, 45) |
| ***b*** = 0.5, 6 months | 0.4 | 37 (36, 39) | 38 (36, 40) |
|  | 0.5 | 44 (43, 46) | 45 (43, 46) |
| ***b*** = 0, 18 months | 0.4 | 2 (0, 4) | 33 (31, 35) |
|  | 0.5 | 6 (4, 8) | 40 (38, 42) |
| ***b*** = 0, 12 months | 0.4 | 14 (12, 16) | 34 (32, 36) |
|  | 0.5 | 17 (14, 18) | 42 (41, 44) |
| ***b*** = 0, 6 months | 0.4 | 29 (28, 31) | 34 (32, 36) |
|  | 0.5 | 34 (32, 35) | 43 (41, 44) |
| ***b*** = 0, 3 months | 0.5 | 44 (43, 45) | 44 (42, 46) |
| ***b*** = 0, at a dose | 0.5 | 45 (44, 46) | 43 (41, 45) |

**Figure S5:**
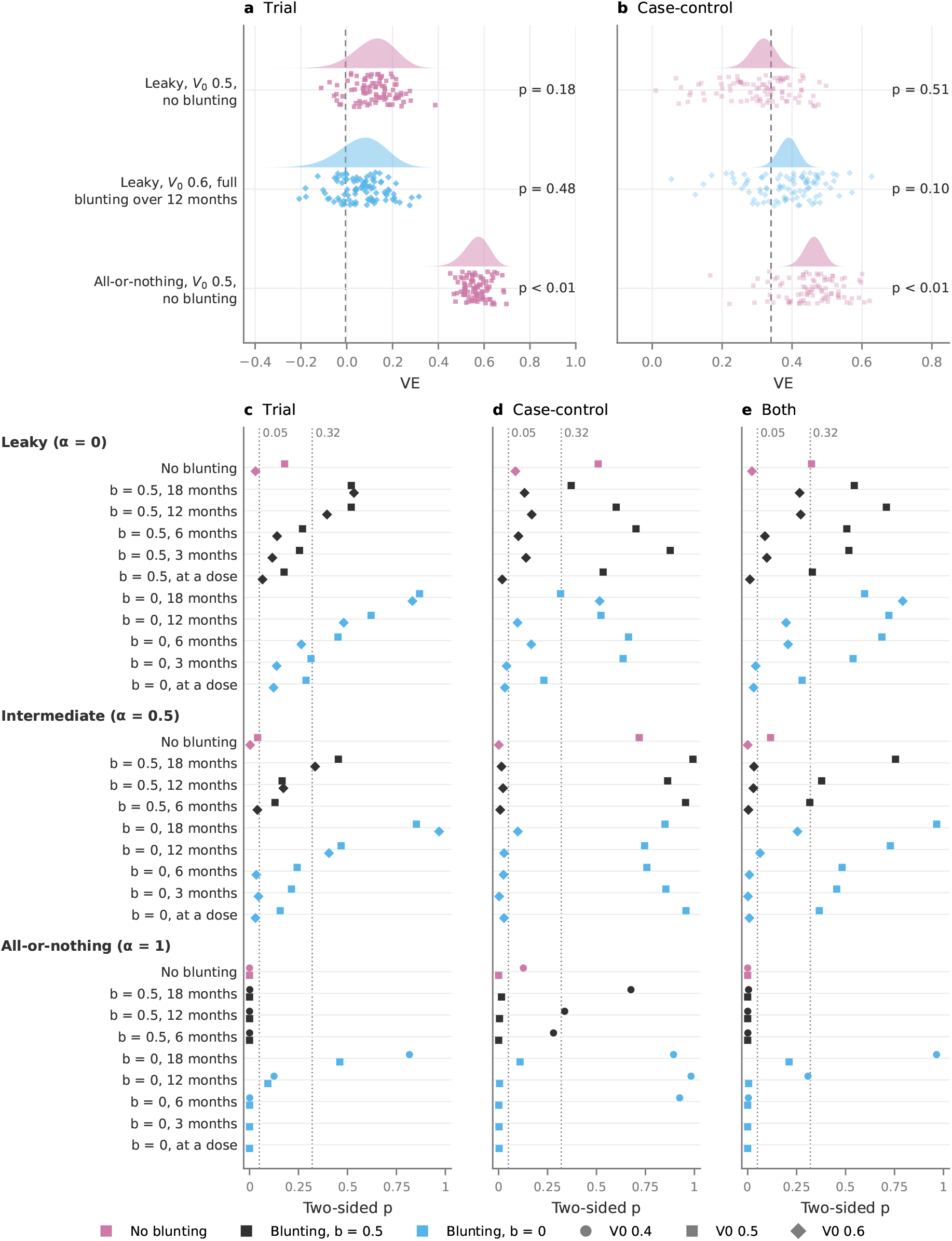
Agreement probabilities in the differential-intensity worlds. How often each vaccine model would give estimates at least as far from its mean estimates as GoGoVax’s^14^ and the pooled case-control estimate.^9^ (a) Trial VE in the 90 simulated GoGoVax-sized trials (dots; 30 runs in each of the three plausible observational settings) for the three vaccine models of Figure S6, with the normal distribution fitted to their log rate ratios (shaded); the dashed line is GoGoVax’s estimate, 0.5%. (b) Case-control VE in the 90 single simulated studies (faint dots) and in simulated meta-analyses of 6–9 runs with the pooled estimate’s precision (shaded); the dashed line is the pooled estimate, 0.34. Numbers give the two-sided probability of the data under each model. (c–e) The two-sided probability for the trial (c), the case-control study (d) and both together (e; *χ*^2^ with 2 degrees of44freedom) for every vaccine model of Figure S4, with rows, colours and marker shapes the same. Dotted lines mark 0.05 and 0.32, the two-sided probabilities at the limits of a 95% and a 68% CI.

**Figure S6:**
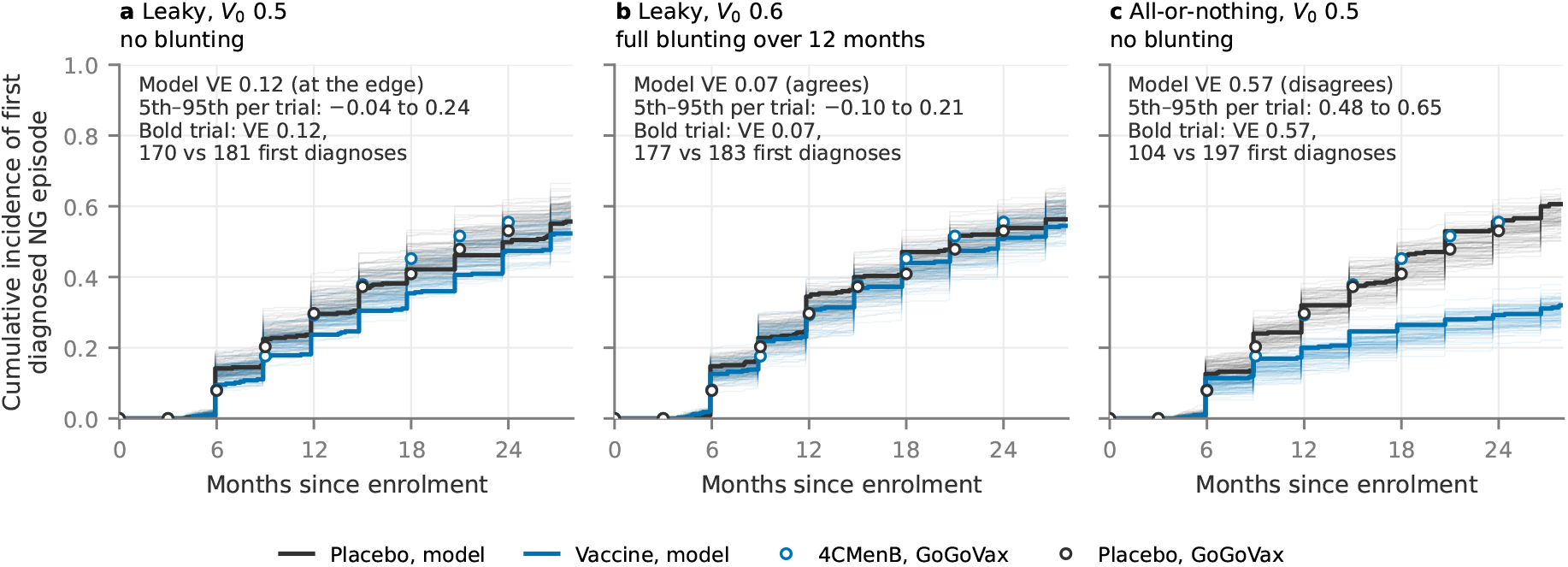
Cumulative incidence of first NG diagnosis in the simulated trials and in GoGoVax, by arm, in the differential-intensity worlds, for (a) leaky protection with *V*_0_ = 0.5 without blunting (at the edge), (b) leaky protection with *V*_0_ = 0.6 and full blunting over the 12 months before dose 1 (agrees), and (c), for contrast, all-or-nothing protection with *V*_0_ = 0.5 without blunting (disagrees). Faint lines show each of the 90 simulated GoGoVax-sized trials (325 per arm; 30 in each of the three plausible observational settings), and bold lines the trial whose VE is nearest the model’s VE, pooled as in Figure 2. Open circles show GoGoVax’s cumulative probability of a first infection in each arm,^14^ reconstructed by the actuarial method from the numbers at risk and cases in its Figure 2. As in GoGoVax, time is counted from enrolment (dose 1), and follow-up starts 28 days after dose 2, at about month 4. The model’s curves rise in steps because its trial visits are exactly three months apart; no one is lost to follow-up, so the cumulative incidence is the share of an arm diagnosed by each day.

## References

1. Tuddenham S, Hamill MM, Ghanem KG. Diagnosis and Treatment of Sexually Transmitted Infections: A Review. JAMA. 2022 Jan;327(2):161–72. Available from: 10.1001/jama.2021.23487.

2. van Hal SJ, Fifer H, Lahra MM. Limitations of Global Surveillance for Neisseria gonorrhoeae Antimicrobial Resistance. Emerg Infect Dis. 2026 Jun;32(6):839–43. Available from: https://wwwnc.cdc.gov/eid/article/32/6/26-0378_article.

3. Seib KL, Grulich AE. Defining the Efficacy of Meningococcal B Vaccines Against Gonococcal Acquisition: The Current Landscape. J Int AIDS Soc. 2026;29(S2):e70153. Available from: https://onlinelibrary.wiley.com/doi/abs/10.1002/jia2.70153.

4. Petousis-Harris H, Paynter J, Morgan J, Saxton P, McArdle B, Goodyear-Smith F, et al. Effectiveness of a group B outer membrane vesicle meningococcal vaccine against gonorrhoea in New Zealand: a retrospective case-control study. Lancet. 2017 Sep;390(10102):1603–10.

5. Abara WE, Bernstein KT, Lewis FMT, Schillinger JA, Feemster K, Pathela P, et al. Effectiveness of a serogroup B outer membrane vesicle meningococcal vaccine against gonorrhoea: a retrospective observational study. Lancet Infect Dis. 2022 Jul;22(7):1021–9.

6. Bruxvoort KJ, Lewnard JA, Chen LH, Tseng HF, Chang J, Veltman J, et al. Prevention of Neisseria gonorrhoeae With Meningococcal B Vaccine: A Matched Cohort Study in Southern California. Clin Infect Dis. 2023 Feb;76(3):e1341–9.

7. Wang B, Giles L, Andraweera P, McMillan M, Almond S, Beazley R, et al. Effectiveness and impact of the 4CMenB vaccine against invasive serogroup B meningococcal disease and gonorrhoea in an infant, child, and adolescent programme: an observational cohort and case-control study. Lancet Infect Dis. 2022;22(7):1011–20.

8. Robison SG, Leman RF. Association of Group B Meningococcal Vaccine Receipt With Reduced Gonorrhea Incidence Among University Students. JAMA Netw Open. 2023 Aug;6(8):e2331742. Available from: https://pmc.ncbi.nlm.nih.gov/articles/PMC10472 183/.

9. Wang B, Mohammed H, Andraweera P, McMillan M, Marshall H. Vaccine effectiveness and impact of meningococcal vaccines against gonococcal infections: A systematic review and meta-analysis. J Infect. 2024 Sep;89(3):106225. Available from: https://www.sciencedirect.com/science/article/pii/S0163445324001592.

10. UK Health Security Agency. Surveillance of targeted 4CMenB vaccination to protect individuals at higher risk of gonorrhoea; 2025. Published 13 November 2025. Accessed 9 November 2026. https://www.gov.uk/government/publications/surveillance-of-4cmenb-vaccination-to-protect-individuals-at-higher-risk-of-gonorrhoea.

11. Molina JM, Bercot B, Assoumou L, Rubenstein E, Algarte-Genin M, Pialoux G, et al. Doxycycline prophylaxis and meningococcal group B vaccine to prevent bacterial sexually transmitted infections in France (ANRS 174 DOXYVAC): a multicentre, open-label, randomised trial with a 2 × 2 factorial design. Lancet Infect Dis. 2024 Oct;24(10):1093–104. Available from: https://www.thelancet.com/journals/laninf/article/PIIS1473-3099(24)00236-6/abstract.

12. Thng C, Eskandari S, Jin F, Hughes I, Grulich AE, O’Sullivan M, et al. Efficacy of the meningococcal vaccine against Neisseria gonorrhoeae: a randomised clinical trial (MenGO). NPJ Vaccines. 2026 Apr;11(1):162. Available from: https://www.nature.com/articles/s41541-026-01422-y.

13. Kwan TH, Chan DPC, Lai CKC, Lam TT, Kwan CK, Lee SS. Efficacy of the 4CMenB vaccine against gonorrhea among men who have sex with men in Hong Kong: A double-blind randomized placebo-controlled trial. Int J Infect Dis. 2026 Sep;170:108926. Available from: https://linkinghub.elsevier.com/retrieve/pii/S1201971226005618.

14. Seib KL, Donovan B, Jin F, Thng C, Yeung B, Comninos N, et al. Meningococcal B Vaccine to Prevent Neisseria gonorrhoeae Infection. N Engl J Med. 2026 Jul;395(4):349–61. Available from: https://www.nejm.org/doi/full/10.1056/NEJMoa2516739.

15. The Lancet Infectious Diseases. GoGoVax dispels hopes for gonorrhoea vaccination. Lancet Infect Dis. 2026 Apr;26(4):329. Editorial.

16. Schnitzer ME. Estimands and estimation of COVID-19 vaccine effectiveness under the test-negative design: Connections to causal inference. Epidemiology. 2022 May;33(3):325–33. Available from: 10.1097/EDE.0000000000001470.

17. Sullivan SG, Tchetgen Tchetgen EJ, Cowling BJ. Theoretical Basis of the Test-Negative Study Design for Assessment of Influenza Vaccine Effectiveness. Am J Epidemiol. 2016 Sep;184(5):345–53.

18. Abara WE, Bernstein KT, Lewis FMT, Pathela P, Islam A, Eberhart M, et al. Healthy vaccinee bias and MenB-FHbp vaccine effectiveness against gonorrhea. Sex Transm Dis. 2023 Jun;50(6):e8–e10. Available from: https://pmc.ncbi.nlm.nih.gov/articles/PMC10175191/.

19. Plummer FA, Chubb H, Simonsen JN, Bosire M, Slaney L, Maclean I, et al. Antibody to Rmp (Outer Membrane Protein 3) Increases Susceptibility to Gonococcal Infection. J Clin Invest. 1993;91(1):339–43.

20. White MT, Griffin JT, Drakeley CJ, Ghani AC. Heterogeneity in malaria exposure and vaccine response: implications for the interpretation of vaccine efficacy trials. Malar J. 2010 Mar;9:82. Available from: https://link.springer.com/article/10.1186/1475-2875-9-82.

21. Edlefsen PT. Leaky Vaccines Protect Highly Exposed Recipients at a Lower Rate: Implications for Vaccine Efficacy Estimation and Sieve Analysis. Comput Math Methods Med. 2014;2014(1):813789. Available from: https://onlinelibrary.wiley.com/doi/abs/10.1155/2014/813789.

22. Hui BB, Padeniya TN, Rebuli N, Gray RT, Wood JG, Donovan B, et al. A Gonococcal Vaccine Has the Potential to Rapidly Reduce the Incidence of Neisseria gonorrhoeae Infection Among Urban Men Who Have Sex With Men. J Infect Dis. 2022 Mar;225(6):983–93. Available from: 10.1093/infdis/jiab581.

23. UK Health Security Agency. Gonorrhoea: the Green Book chapter; 2025. In: Immunisation against infectious disease (the Green Book). Version of 16 June 2025. Accessed 9 November 2026. https://www.gov.uk/government/publications/gonorrhoea-the-green-book-chapter.

24. Ogaz D, Mullen D, Habib Meriggi G, Hoerst C, Reid D, Jayes D, et al. New era in STI prevention: early adoption of doxyPEP and 4CMenB vaccination among a community sample of men and gender-diverse people having sex with men in England—findings from a large, cross-sectional survey in 2025. Sex Transm Infect. 2026. Published online 24 July 2026.

25. Dutt K, Chow EPF, Huffam S, Klassen K, Fairley CK, Bradshaw CS, et al. High Prevalence of Rectal Gonorrhoea among Men Reporting Contact with Men with Gonorrhoea: Implications for Epidemiological Treatment. BMC Public Health. 2015 Jul;15(1):658.

26. Dolling DI, Desai M, McOwan A, Gilson R, Clarke A, Fisher M, et al. An analysis of baseline data from the PROUD study: an open-label randomised trial of pre-exposure prophylaxis. Trials. 2016 Mar;17:163.

27. Mercer CH, Jones KG, Johnson AM, Lewis R, Mitchell KR, Gravningen K, et al. How can we objectively categorise partnership type? A novel classification of population survey data to inform epidemiological research and clinical practice. Sex Transm Infect. 2017;93(2):129–36.

28. Althaus CL, Choisy M, Alizon S, CSF group. How sex acts scale with the number of sex partners: evidence from *Chlamydia trachomatis* data and implications for control. PeerJ PrePrints. 2015;3:e940v2. Preprint.

29. Wang B, Giles L, Andraweera P, McMillan M, Beazley R, Almond S, et al. Long-Term Protection Against Invasive Meningococcal B Disease and Gonococcal Infection 5 Years After Implementation of Funded Childhood and Adolescent 4CMenB Vaccination Program in South Australia: An Observational Cohort and Case-Control Study. Clin Infect Dis. 2025 Jul;81(4):e202–10. Available from: https://pmc.ncbi.nlm.nih.gov/articles/PMC12596398/.

30. National Institute of Allergy and Infectious Diseases. Safety and Efficacy Study of Meningococcal Group B Vaccine rMenB+OMV NZ (Bexsero) to Prevent Gonococcal Infection; 2020. MAGI trial (DMID 19-0004/HPTN 107). First posted 16 April 2020; last updated 24 July 2026; accessed 15 November 2026. ClinicalTrials.gov identifier NCT04350138. Available from: https://clinicaltrials.gov/study/NCT04350138.

31. University of Washington. BIYELA – Bexsero Immunisation in Young Women in Africa; 2024. First posted 6 June 2024; last updated 10 July 2025; accessed 15 November 2026. ClinicalTrials.gov identifier NCT06446752. Available from: https://clinicaltrials.gov/study/NCT06446752.

32. Kretzschmar M, van Duynhoven YT, Severijnen AJ. Modeling prevention strategies for gonorrhea and Chlamydia using stochastic network simulations. Am J Epidemiol. 1996 Aug;144(3):306–17.

33. Ghani AC, Swinton J, Garnett GP. The role of sexual partnership networks in the epidemiology of gonorrhea. Sex Transm Dis. 1997 Jan;24(1):45–56.

34. Jenness SM, Weiss KM, Goodreau SM, Gift T, Chesson H, Hoover KW, et al. Incidence of Gonorrhea and Chlamydia Following Human Immunodeficiency Virus Preexposure Prophylaxis Among Men Who Have Sex With Men: A Modeling Study. Clin Infect Dis. 2017 Sep;65(5):712–8. Available from: 10.1093/cid/cix439.

35. Padeniya TN, Hui BB, Wood JG, Regan DG, Seib KL. Review of mathematical models of Neisseria gonorrhoeae vaccine impact: implications for vaccine development. Vaccine. 2024 Jul;42 (suppl 1)(19):S70–81.

36. Prah P, Hickson F, Bonell C, McDaid LM, Johnson AM, Wayal S, et al. Men who have sex with men in Great Britain: comparing methods and estimates from probability and convenience sample surveys. Sex Transm Infect. 2016;92(6):455–63.

37. Clifton S, Fuller E, Philo D. National Survey of Sexual Attitudes and Lifestyles (Natsal-3): Reference Tables. London: NatCen Social Research, on behalf of the Natsal team; n.d.. Table 29: Number of opposite-sex partners in last year, by age and sex. Accessed 4 November 2026. Available from: https://www.natsal.ac.uk.

38. Barbee LA, Soge OO, Khosropour CM, Haglund M, Yeung W, Hughes J, et al. The Duration of Pharyngeal Gonorrhea: A Natural History Study. Clin Infect Dis. 2021;73(4):575–82.

39. Chow EPF, Camilleri S, Ward C, Huffam S, Chen MY, Bradshaw CS, et al. Duration of gonorrhoea and chlamydia infection at the pharynx and rectum among men who have sex with men: a systematic review. Sex Health. 2016;13(3):199–204.

40. Davies EP, Tsuboi M, Evans J, Rowley J, Korenromp EL, Clayton T, et al. A global meta-analysis of gonorrhoea and chlamydia prevalence among men who have sex with men from 2000 to 2022. Int J STD AIDS. 2025;36(8):611–21.

41. Sonnenberg P, Clifton S, Beddows S, Field N, Soldan K, Tanton C, et al. Prevalence, risk factors, and uptake of interventions for sexually transmitted infections in Britain: findings from the National Surveys of Sexual Attitudes and Lifestyles (Natsal). Lancet. 2013;382(9907):1795–806.

42. Erens B, Phelps A, Clifton S, Mercer CH, Tanton C, Hussey D, et al. Methodology of the third British National Survey of Sexual Attitudes and Lifestyles (Natsal-3). Sex Transm Infect. 2014 Mar;90(2):84–9. Available from: https://sti.bmj.com/content/90/2/84.

43. Whittles LK, Didelot X, White PJ. Public health impact and cost-effectiveness of gonorrhoea vaccination: an integrated transmission-dynamic health-economic modelling analysis. Lancet Infect Dis. 2022 Jul;22(7):1030–41. Available from: https://www.thelancet.com/journals/laninf/article/PIIS1473-3099(21)00744-1/fulltext.

44. Midtbø JE, Rø G, Gran JM, Kristoffersen AB, Meijerink H, LeBlanc M. Vaccine Effectiveness Is Not a Model Parameter, Even in a Low-Attack-Rate Trial; 2026. Manuscript in preparation.

45. Wang B, Giles L, Andraweera P, McMillan M, Almond S, Beazley R, et al. 4CMenB sustained vaccine effectiveness against invasive meningococcal B disease and gonorrhoea at three years post programme implementation. J Infect. 2023;87(2):95–102.

46. Raccagni AR, Galli L, Spagnuolo V, Bruzzesi E, Muccini C, Bossolasco S, et al. Meningococcus B Vaccination Effectiveness Against Neisseria gonorrhoeae Infection in People Living With HIV: A Case-Control Study. Sex Transm Dis. 2023;50(5):247–51.

47. Abara WE, Modaressi S, Fireman B, Klein NP, Layefsky E, Goddard K, et al. Effectiveness of a serogroup B meningococcal vaccine against gonorrhea: A retrospective study. Vaccine. 2024;42(26):126312.

48. McMillan M, Wang B, Krause V, Greenwood-Smith B, Webby R, Chen J, et al. A Multidesign Study of 4CMenB Vaccine Effectiveness and Impact on Meningococcal Carriage and Gonorrhea in Northern Territory Adolescents and Young People. Open Forum Infect Dis. 2026;13(8):ofag409.

49. UK Health Security Agency. Sexually transmitted infections quarterly report, England, June 2026; 2026. Provisional data. Accessed 9 November 2026. https://www.gov.uk/government/publications/sti-quarterly-surveillance-reports-provisional-data/sexually-transmitted-infections-quarterly-report-england-june-2026.

50. UK Health Security Agency. Sexually transmitted infections quarterly report, England: provisional data, September 2026; 2026. Accessed 9 November 2026. https://www.gov.uk/government/publications/sti-quarterly-surveillance-reports-provisional-data/sexually-transmitted-infections-quarterly-report-england-provisional-data-september-2026.

